# Long-term risk of post-acute sequelae among adults following SARS-CoV-2 or influenza virus infection: a retrospective cohort study in a large, integrated healthcare system

**DOI:** 10.1101/2025.05.30.25328674

**Authors:** Joseph A. Lewnard, Debbie E. Malden, Vennis Hong, Jessica Skela, Leora R. Feldstein, Sharon Saydah, Iris Anne C. Reyes, Rulin Hechter, Lina S. Sy, Bradley K. Ackerson, Sara Y. Tartof

**Affiliations:** School of Public Health, University of California, Berkeley, 2121 Berkeley Way, Berkeley, California 94720, United States; Kaiser Permanente Southern California Department of Research & Evaluation, 100 S. Los Robles, Pasadena, California 91101, United States; Centers for Disease Control & Prevention, 1600 Clifton Rd. NE, Atlanta, Georgia 30333, United States; Department of Health Systems Science, Kaiser Permanente Bernard J. Tyson School of Medicine, 98 S. Los Robles, Pasadena, California 91101, United States

## Abstract

**Background:** The comparative risk of post-acute sequelae (PAS) associated with SARS-CoV-2 and influenza virus infection remains unclear.

**Methods:** We undertook a retrospective cohort study within the Kaiser Permanente Southern California healthcare system of COVID-19 and influenza cases who received acute respiratory illness (ARI) diagnoses in virtual, outpatient, or inpatient settings between 1 September, 2022 and 31 December, 2023. We monitored PAS-associated healthcare utilization across all settings through 180 days after index ARI diagnoses. We estimated adjusted hazard ratios (aHRs) comparing COVID-19 cases to influenza cases, weighting to account for cases’ probability of retention in follow-up and infection with SARS-CoV-2 or influenza virus at the index ARI episode.

**Results:** Analyses included 74,738 COVID-19 cases and 18,790 influenza cases, among whom 35,835 (38.3%), 26,579 (28.4%), 23,388 (25.0%), and 7,726 (8.3%) received care for their index ARI episodes in virtual, ambulatory, emergency department, and inpatient settings, respectively. Risk of PAS diagnoses in any clinical setting was similar among COVID-19 and influenza cases (aHR=1.04 [95% confidence interval: 0.99-1.09] and aHR=1.01 [0.97-1.06] 31-90 and 91-180 days after index, respectively). However, COVID-19 cases experienced higher risk of severe PAS conditions necessitating inpatient care (aHR=1.31 [1.07-1.59] and aHR=1.24 [1.03-1.49] 31-90 and 91-180 days after index, respectively). This heightened risk of severe PAS following COVID-19 was concentrated among patients who required inpatient admission at their index episode.

**Conclusions:** PAS outcomes occur with similar frequency among non-severe COVID-19 cases and influenza cases. However, PAS among COVID-19 cases are more likely to require hospital admission than PAS among influenza cases.

**Key points:**

- Non-severe acute respiratory illnesses involving SARS-CoV-2 and influenza virus are associated with similar risk of post-acute sequelae (PAS).
- Severe COVID-19 cases experience greater risk of PAS than severe influenza cases.
- PAS following COVID-19 are more likely to necessitate hospital admission.

## INTRODUCTION

Acute respiratory infections are associated with post-acute sequelae (PAS), a complex constellation of symptoms which may involve multiple organ systems. Longitudinal studies have estimated that as many as 30% of individuals may experience at least one post-acute symptom or PAS condition ≥30 days after SARS-CoV-2 infection.^1–3^ Some studies have demonstrated persistent presentation of PAS for up to 3 years following the acute stages of COVID-19.^4^ Although less extensively studied, PAS including neurological conditions^5,6^ and chronic fatigue^7^ have also been documented after influenza infection, with reports of severe or prolonged post-influenza conditions dating to the 1918 pandemic.^8^ However, it remains unclear whether SARS-CoV-2 and influenza infections pose comparable risk for PAS, and whether the clinical phenotypes of PAS associated with SARS-CoV-2 and influenza are distinct.

Several comparative studies^9,10^ have reported greater risk of PAS after COVID-19 in comparison to influenza, albeit inconsistently.^11^ However, a key limitation of these studies has been their restriction to hospitalized COVID-19 and influenza patients, as PAS also affect patients whose preceding viral infections were not severe.^12^ Persons hospitalized for SARS-CoV-2 and influenza are not representative of all people infected with these viruses, and may experience different PAS outcomes in comparison to non-hospitalized patients. We therefore aimed to assess the risk of PAS following COVID-19 and influenza among adults, including a broad sample of individuals whose initial illness varied in clinical severity. We compared PAS outcomes measured based on subsequent healthcare utilization across settings spanning virtual to inpatient care in analyses that applied stringent correction for bias due to confounding and retention.

## METHODS

### Setting

We analyzed data collected from members of Kaiser Permanente Southern California (KPSC), an integrated health-care organization comprised of 16 hospitals and 226 medical offices providing care to >4.8 million members across Southern California. Members reflect the socioeconomic and racial and ethnic diversity of the area’s population.^13,14^ All aspects of clinical care received at KPSC facilities, including diagnoses, clinical notes, laboratory tests, and prescriptions, are linked by a unique identifier in patients’ electronic health records (EHRs). Insurance claims submitted for reimbursement capture care received from external healthcare providers, enabling near-complete capture of healthcare receipt among members.

### Study design

We conducted a retrospective cohort study of all KPSC members aged ≥18 years with a positive molecular test for SARS-CoV-2 or influenza between September 1, 2022 and December 31, 2023. We restricted analyses to individuals with accompanying acute respiratory illness (ARI) diagnoses in any clinical setting between 7 days before or after the positive test.

Eligible participants had ≥1 year of continuous KPSC membership (allowing 45-day enrollment gap) before the positive test date to enable accurate capture of comorbid conditions and healthcare utilization. We limited analyses to the first documented ARI diagnosis associated with a positive SARS-CoV-2 or influenza test result in any 180-day period for each individual (index ARI episode). Multiple index episodes were included from the same individual if these episodes occurred ≥180 days apart. We excluded SARS-CoV-2 infections identified between 30 days before and 14 days after individuals received any COVID-19 vaccine dose, and influenza infections identified 30 days before to 14 days after individuals received any influenza vaccine, so that participants’ recorded vaccination status at time of infection reflected doses from which they could be expected to have mounted a response. We excluded patients with missing information on age or sex to enable adjustment for confounding.

### Exposures

The primary exposure was the virus detected at each index ARI episode; we defined exposure groups as COVID-19 cases and influenza cases. We stratified further stratified exposure groups based on respiratory viral season (October, 2022–September, 2023, and October–December, 2023), differing viral lineages (influenza type A or B, where available from testing data, and putative SARS-CoV-2 variant based on dominant lineages circulating during the respective time periods^15^). Additionally, we distinguished index ARI episodes according to the highest-acuity care setting in which individuals received ARI diagnoses. In order from lowest to highest acuity, strata included virtual (online/telehealth) settings, ambulatory (outpatient/urgent care) settings, emergency departments, and inpatient (hospital) facilities.

### Outcomes

We defined PAS outcomes as diagnoses occurring 31-180 days after cases’ index ARI episodes associated with a confirmed SARS-CoV-2 or influenza virus infection. We used prespecified ICD-10 diagnoses codes categorized into 10 disease categories with similar mechanisms and body systems affected (cardiopulmonary, hemolytic, respiratory, musculoskeletal, renal, gastrointestinal, neurological, skin, endocrine, and mental health conditions; **Table S1**). Although there is no universal case definition for PAS, this definition included published US Centers for Disease Control criteria for Post-COVID Conditions,^2^ augmented with definitions from other EHR-based studies^16–19^ We conducted separate analyses for PAS affecting each disease category, and for a composite outcome of any PAS. Consistent with our classification for index ARI diagnoses, we took the highest-acuity care setting where PAS diagnoses were assigned in each follow-up period as an indicator of severity..

We censored observations at death, disenrollment, or end of study period, whichever occurred first. We stratified follow-up into proximal (31-90 days) and distal (91-180 days) periods following each index ARI episode.

### Statistical analysis

We measured associations via the adjusted hazard ratio (aHR) for time to first PAS diagnosis among COVID-19 cases versus influenza cases. Consistent with target-trial emulation frameworks,^20^ we used a two-stage weighting approach to mitigate selection bias (due to differential depletion of susceptibles prior to follow-up initiation) and confounding. First, to address differential censoring prior to the beginning of each follow-up interval among COVID-19 cases and influenza cases, we generated inverse probability of censoring weights (IPCWs) addressing each individual’s probability of remaining enrolled through the beginning of each 30-day period after index.^21^ We estimated IPCWs via the Breslow estimator for survival curves from fitted Cox proportional hazards models with death or disenrollment 0-180 days after cases’ index date as the outcome. Second, to account for differences in characteristics of COVID-19 cases and influenza cases, we also generated inverse probability of treatment (exposure) weights (IPTWs), addressing each individual’s probability of an index infection with SARS-CoV-2 or influenza. We computed stabilized IPTWs via logistic regression models defining infecting virus as the outcome.^22^ All weighting models accounted for individuals’ age group, sex, highest-acuity care setting for the index ARI episode, race/ethnicity, Charlson comorbidity index, history of diagnoses in each PAS disease category, history of depression, cigarette smoking, prior-year healthcare utilization (across ambulatory, emergency department, and inpatient settings), body mass index, COVID-19 and seasonal influenza vaccination status, calendar month of the index episode, and neighborhood deprivation index.^23^ We list categorizations for continuous variables in **Table 1**.

**Table 1:**
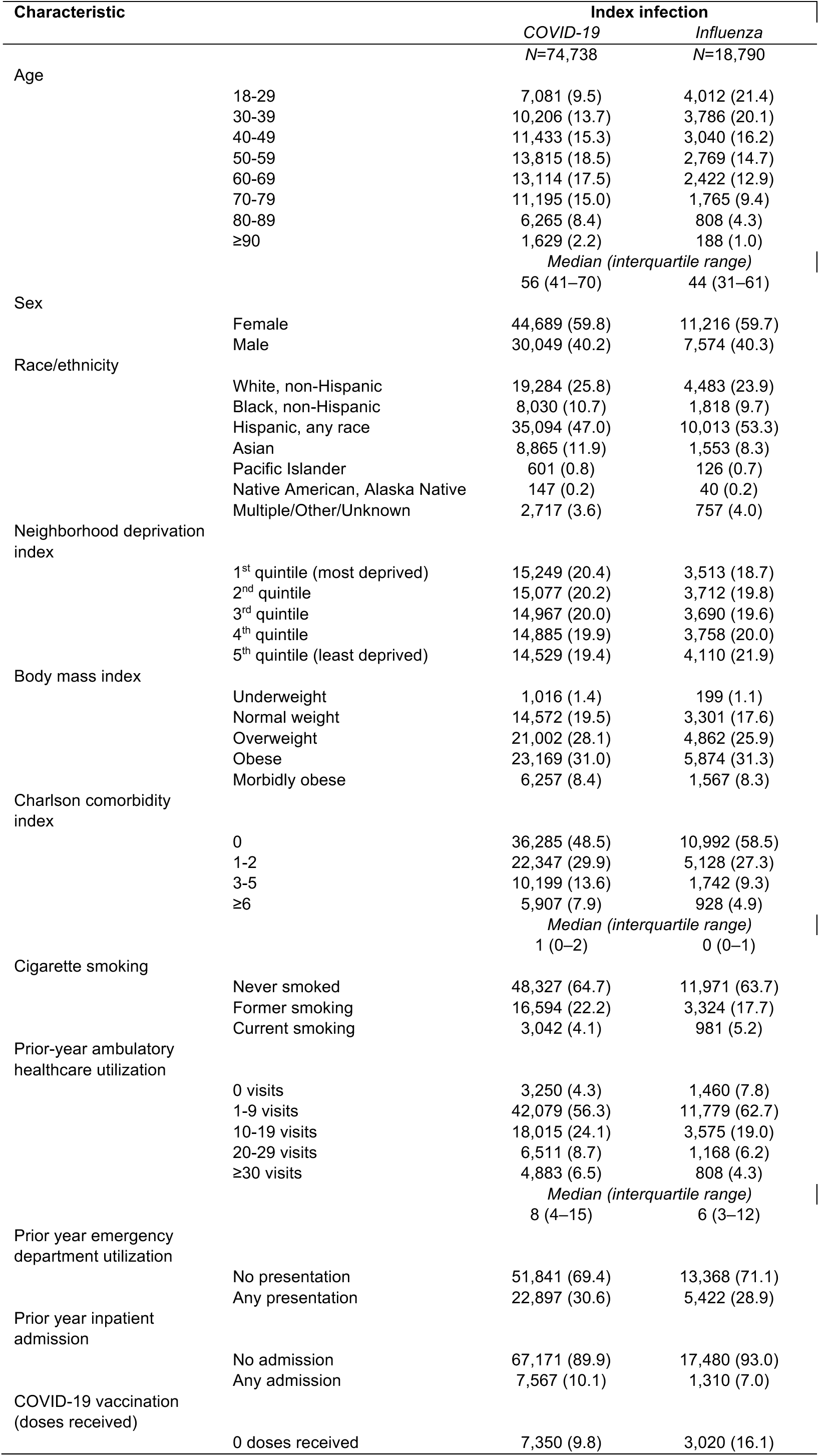
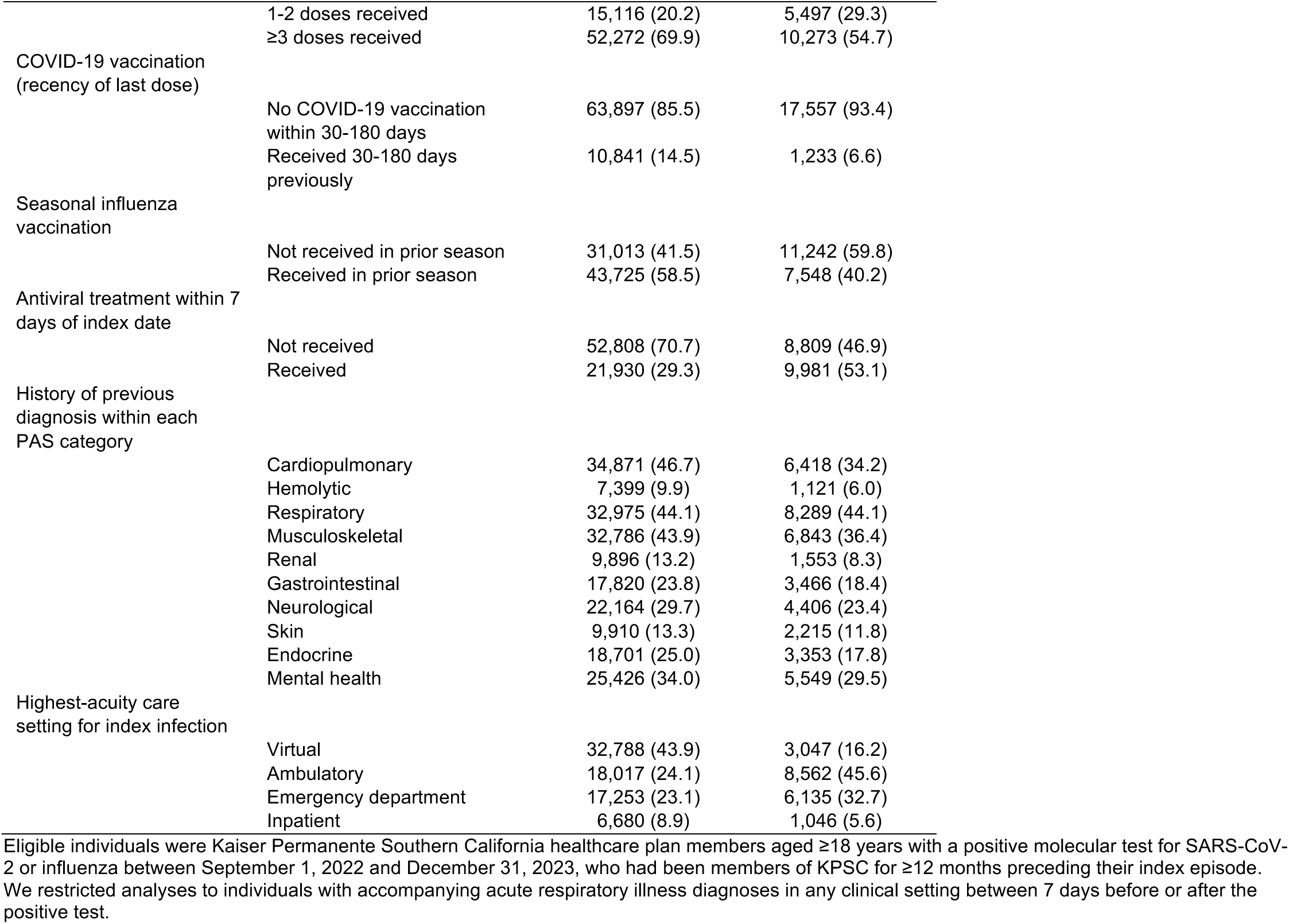
Characteristics of COVID-19 cases and influenza cases.

We estimated aHRs using Cox proportional hazards models, defining distinct observations for each 30-day period (31-60, 61-90, 91-120, 121-150, 151-180 days) after the index date to update IPCWs. Weights for each follow-up interval were the product of the individual’s (time-varying) IPCW and the individual’s (time-invariant) IPTW for infection with the identified virus.^24^ We generated doubly-robust aHR estimates by including covariates used in the weighting models in analysis models, and used the sandwich variance estimator to account for repeated observations across multiple periods for each individual. We fit separate models for follow-up periods 31-90 days and 91-180 days after index. We verified the proportional hazards assumption by testing for non-zero slopes of Schoenfeld residuals from fitted Cox proportional hazards models.^25^ We considered differences in risk to be statistically meaningful if 95% confidence intervals around aHR estimates excluded the null value.

We conducted subgroup analyses restricted to individuals with or without history of prior diagnoses corresponding to PAS conditions within each organ system category in the preceding year. Among persons with such history, we interpreted PAS outcomes as exacerbations of pre-existing conditions, such that aHR estimates conveyed the differences in risk of post-viral exacerbations among COVID-19 cases and influenza cases. Among persons without history of such diagnoses, we interpreted PAS outcomes as new-onset illness, such that aHR estimates conveyed the differential risk of new-onset PAS among COVID-19 cases and influenza cases.

Last, we conducted subgroup analyses stratified by age, sex, severity of the index ARI episode, and receipt of antivirals and vaccines. We defined antiviral receipt for the index infection as dispenses of molnupiravir or nirmatrelvir-ritonavir (for COVID-19 cases) and oseltamivir, zanamivir, peramivir, or baloxavir (for influenza cases), occurring within 7 days before or after index. We defined up-to-date influenza vaccination status as receipt of seasonal influenza vaccination within 15-180 days before index. Based on US vaccination recommendations during the study period^26^ and estimated durations of protection against SARS-CoV-2 infection after vaccination,^27–29^ we defined up-to-date COVID-19 vaccination as receipt of ≥3 COVID-19 vaccine doses, including one dose within 15-180 days before index.

### Ethics

This study was reviewed and approved by the KPSC institutional review board with a waiver of informed consent and was conducted consistent with federal law and CDC policy (45 C.F.R. part 46.101(c); 21 C.F.R. part 56).

## RESULTS

Analyses included 74,738 eligible COVID-19 cases and 18,790 eligible influenza cases. In comparison to influenza cases, COVID-19 cases were older, had higher Charlson comorbidity index scores, and had higher outpatient healthcare utilization in the year prior to index date as well as greater likelihood of hospital admission in the prior year (**Table 1**). Additionally, COVID-19 cases had higher likelihood of receiving care in either inpatient settings or in virtual settings only.

Overall, 97.3% and 95.3% of COVID-19 cases were retained in follow-up through 31 and 91 days after their index date, respectively (72,745 and 71,238 of 74,738, respectively), as were 97.9% and 95.7% of influenza cases (18,395 and 17,990 of 18,790, respectively; **Table S2**). Within 31-90 days after index dates, 45.2% of COVID-19 cases and 38.9% of influenza cases received PAS diagnoses, and within 91-180 days after index dates, 52.8% of COVID-19 cases and 44.0% of influenza cases received PAS diagnoses. Cardiopulmonary PAS were the most common outcome among both COVID-19 cases and influenza cases (23.9% and 16.1% of cases, respectively).

Absolute differences in weighted incidence of PAS diagnoses by virus were modest in analyses adjusting for individuals’ probability of infection with each virus and censoring (**Table 2**). Within 31-90 days after index, weighted incidence rates of PAS diagnoses per 100 person-months at risk were 26.5 (95% confidence interval: 26.2-26.8) among COVID-19 cases and 25.9 (25.4-26.5) among influenza cases; within 91-180 days after index, weighted incidence rates were 23.0 (22.7-23.2) and 22.6 (22.1-23.0) per 100 person-months among COVID-19 and influenza cases, respectively. In terms of relative (proportional) differences in weighted PAS incidence rates comparing COVID-19 cases and influenza cases, differentiation was most apparent for PAS diagnoses in inpatient settings. We observed 1.5 (1.4-1.5) and 1.2 (1.1-1.4) inpatient PAS diagnoses per 100 person-months among COVID-19 and influenza cases, respectively, 31-90 days after index, and 1.1 (1.0-1.1) and 0.9 (0.8-1.0) inpatient PAS diagnoses per 100 person-months among COVID-19 and influenza cases, respectively, 91-180 days after index.

**Table 2:**
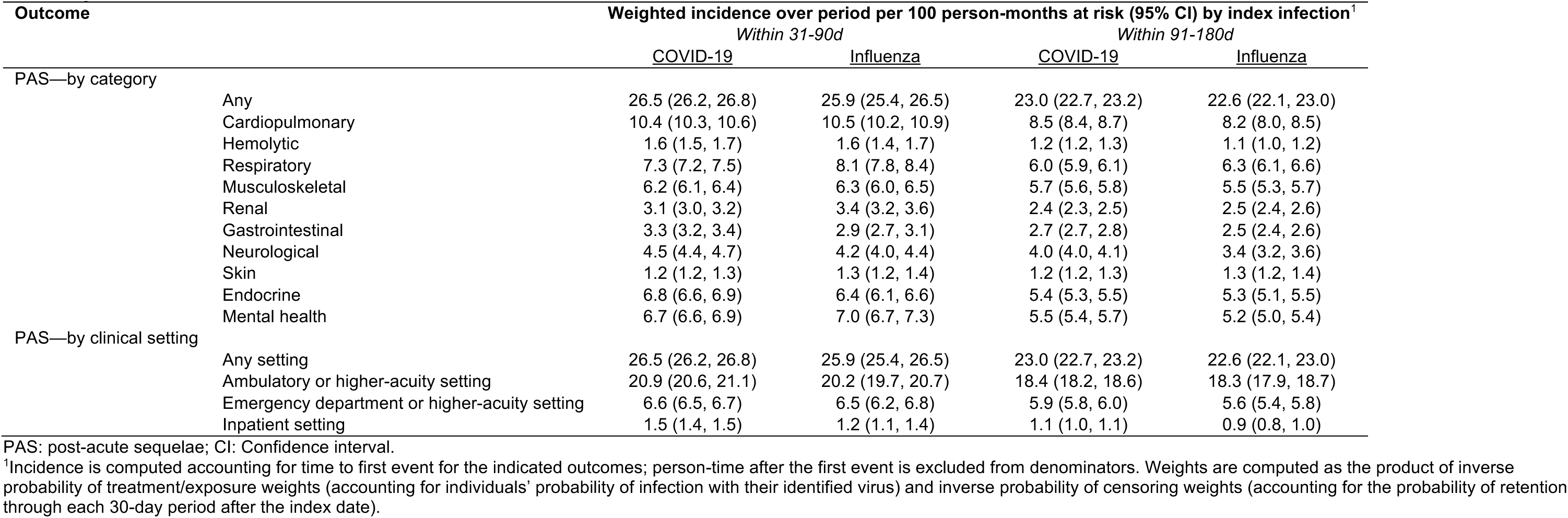
Rates of post-acute sequelae managed in various clinical settings, adjusted via weighting to address loss-to-follow up and differences among cases by index infection.

Using doubly-robust Cox proportional hazards models, we did not identify appreciable differences in risk of PAS outcomes among COVID-19 cases and influenza cases either 31-90 days or 91-180 days after cases’ index dates (aHR=1.04 [95% confidence interval: 0.99-1.09] and aHR=1.01 [0.97-1.06] at 31-90 and 91-180 days after index, respectively, for COVID-19 versus influenza cases; **Table 3**). However, COVID-19 cases had 31% (7-59%) and 24% (3-49%) higher risk than influenza cases of receiving PAS diagnoses in inpatient settings 31-90 days and 91-180 days after index in comparison to influenza cases, respectively. This higher risk of inpatient PAS diagnoses among COVID-19 cases versus influenza cases was apparent in the 31-90 day follow-up period for cardiopulmonary, hemolytic, respiratory, gastrointestinal, neurological, skin, endocrine, and mental health PAS categories. We also identified higher risk of cardiopulmonary, gastrointestinal, neurological, skin, endocrine, and mental health PAS diagnoses in lower-acuity settings among COVID-19 cases than influenza cases; point estimates suggested a smaller difference in risk among COVID-19 versus influenza cases for PAS diagnoses within virtual, ambulatory, and emergency department settings than within inpatient settings for each of these syndromic categories. In comparison to influenza cases, COVID-19 cases also experienced higher risk of cardiopulmonary, gastrointestinal, neurological, and mental health PAS diagnoses in inpatient settings 91-180 days after index.

**Table 3:**
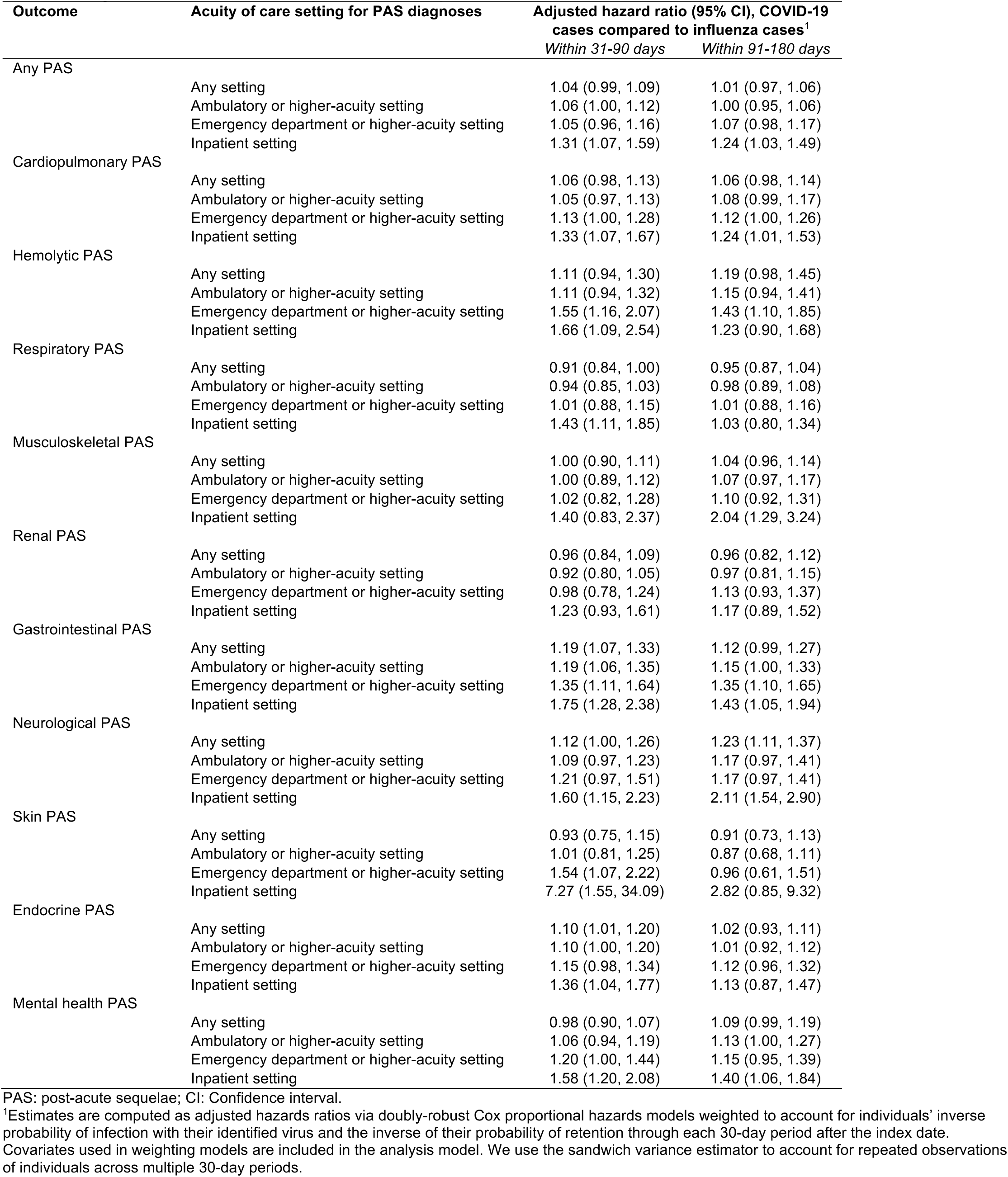
Adjusted hazard ratios of post-acute sequelae.

Similarly, differences between COVID-19 cases and influenza cases in risk of PAS exacerbations of pre-existing conditions were mainly evident for diagnoses in inpatient settings (**Table 4**). Within 31-90 days after index, COVID-19 cases with pre-existing cardiopulmonary, hemolytic, gastrointestinal, neurological, endocrine, and mental health diagnoses experienced 32-97% greater risk of exacerbations resulting in inpatient diagnoses for these conditions than influenza cases with pre-existing diagnoses within the same syndromic categories. Within 91-180 days after index, COVID-19 cases experienced 41-216% greater risk of exacerbations leading to inpatient diagnoses of pre-existing musculoskeletal, gastrointestinal, neurological, and mental health conditions in comparison to influenza cases. For new-onset PAS diagnoses, COVID-19 experienced greater risk than influenza cases of respiratory diagnoses in inpatient settings 31-90 days after index, and of renal conditions in inpatient settings 91-180 days after index. However, estimated aHRs had greater uncertainty for comparisons of new-onset PAS diagnoses than for PAS exacerbations, reflecting the low frequency of new-onset PAS diagnoses compared to PAS exacerbations (**Table S3**).

**Table 4:**
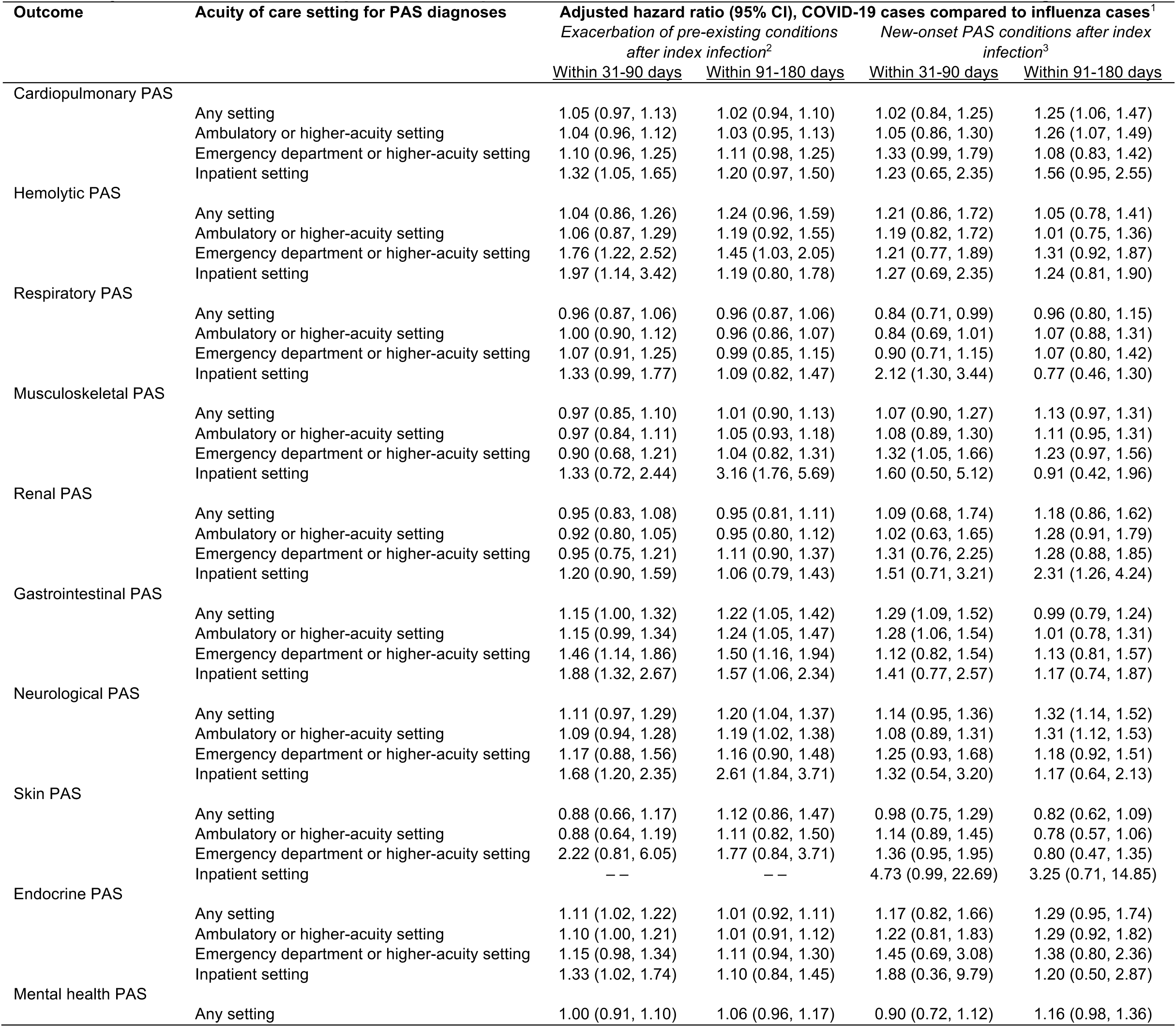
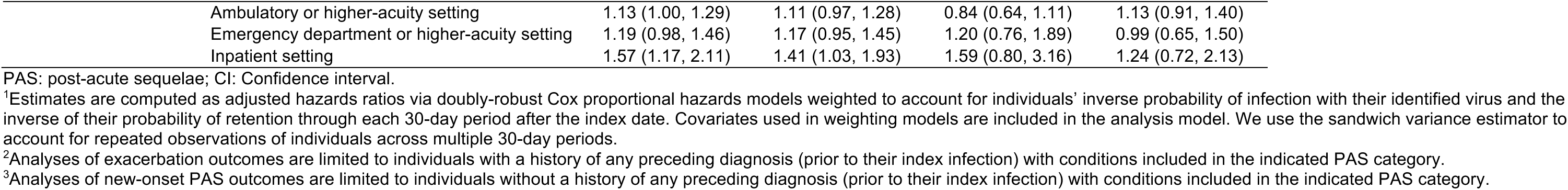
Adjusted hazards ratios of post-acute sequelae as new-onset conditions or exacerbations of existing conditions.

Higher risk of inpatient PAS diagnoses among COVID-19 cases compared to influenza cases was primarily apparent among patients whose index ARI episodes necessitated inpatient admission (**Table 5**). Compared to influenza cases who received ARI diagnoses in inpatient settings, COVID-19 cases who received ARI diagnoses in inpatient settings had 73% (30-130%) and 49% (9-106%) higher risk of inpatient PAS diagnoses 31-90 and 91-180 days after index, respectively. Evidence of increased risk of PAS diagnoses among COVID-19 cases was inconsistent among patients whose index ARI episodes were managed in lower-acuity care settings. Differences in risk of inpatient PAS diagnoses among COVID-19 cases and influenza cases were apparent only among cases who did not receive antiviral treatment (aHR=2.09 [1.45-3.03] for untreated COVID-19 cases versus untreated influenza cases within 31-90 days after index). We did not identify differences in risk of inpatient PAS diagnoses among COVID-19 cases and influenza cases who were up-to-date with COVID-19 and influenza vaccines, respectively, at the time of their index ARI episode. Differences in risk of inpatient-diagnosed PAS conditions among COVID-19 cases versus influenza cases were more strongly apparent among female cases than male cases; additionally, differences in PAS risk among COVID-19 cases versus influenza cases appeared within multiple age groups.

**Table 5:**
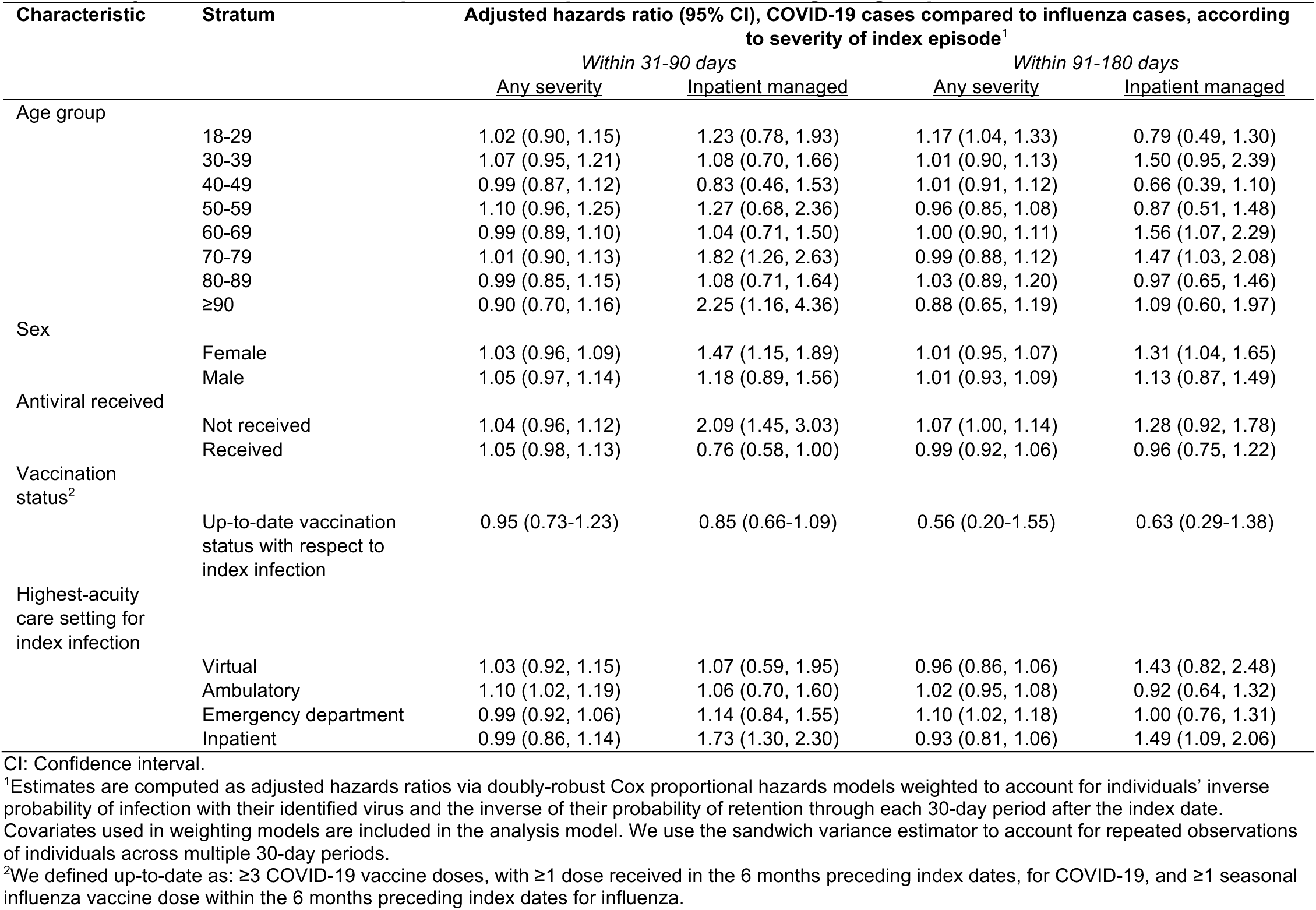
Adjusted hazard ratios of post-acute sequelae across differing subgroups.

Our finding that COVID-19 cases experienced greater risk than influenza cases of severe PAS necessitating hospital admission held within analyses disaggregated by season (2022-23 and 2023-24), SARS-CoV-2 variant period (BA.4/BA.5, XBB/XBB.1.5, and BA.2.86/JN.1), and influenza comparator virus (A or B), although these analyses encountered lower statistical power than primary analyses (**Table S4**). Consistent with our primary analyses, we did not identify appreciable differences in risk of PAS outcomes that included diagnoses in lower-acuity settings within most subgroups (**Table 5**; **Table S4**).

## DISCUSSION

Within our study, COVID-19 cases and influenza cases experienced similar risk of PAS diagnoses up to 180 days after their index ARI episode. The primary distinguishing characteristic of PAS following COVID-19 verus influenza was that COVID-19 cases experienced higher risk of severe PAS necessitating inpatient admission. Further, these differences in risk were dependent on the severity of cases’ index infection. Whereas risk of severe PAS was comparable among COVID-19 cases and influenza cases who received care for their index episode in virtual, ambulatory, or emergency department settings, COVID-19 cases hospitalized for their index infection had higher risk of severe PAS than influenza cases who were hospitalized for their index infection.

Differences in PAS risk were attenuated in strata of patients who received antiviral treatment or whose vaccination status was up-to-date against their infecting virus. Our findings demonstrate that COVID-19 cases and influenza cases experience similar risk of PAS, although the spectrum of PAS occurring after COVID-19 includes more severe manifestations than PAS occurring after influenza. This heightened risk of severe PAS is concentrated among patients who experienced severe COVID-19, and may be attenuated by vaccination or antiviral treatment.

Cardiopulmonary manifestations accounted for the greatest share of all PAS outcomes observed among both COVID-19 and influenza cases, and have been widely documented as sequelae of respiratory virus infections, not limited to SARS-CoV-2 and influenza.^8–11,30–32^ Although less common, severe PAS outcomes involving hemolytic, gastrointestinal, neurological, and skin conditions showed the greatest relative degree of elevation among COVID-19 cases compared to influenza cases within 31-90 days after index. Within 91-180 days after index, severe PAS outcomes involving musculoskeletal, gastrointestinal, neurological, and skin conditions showed the greatest relative degree of elevation among COVID-19 cases.

Among individuals with pre-existing conditions corresponding to each PAS category, COVID-19 cases experienced higher risk than influenza cases of exacerbations leading to inpatient PAS diagnoses. Point estimates of aHRs were consistent with the hypothesis that COVID-19 cases also experienced greater risk of new-onset severe PAS in comparison to influenza cases. However, given the low incidence of severe PAS among individuals without pre-existing conditions, such differences were uncertain from a statistical perspective and may not be clinically meaningful.

Comprehensive capture of healthcare interactions in an integrated system enabled us to represent a broad severity spectrum of index ARI episodes and subsequent PAS in comparison to prior studies which addressed only hospitalized COVID-19 cases and influenza cases.^9,10^ Moreover, access to detailed information on cases’ demographics, comorbidities, and prior patterns of healthcare utilization enabled us to adjust rigorously for differences in characteristics of COVID-19 cases and influenza cases, as well as time-dependent sources of bias not addressed in prior studies. The diverse demographic and clinical characteristics of the enrolled population furthermore support external generalizability of our results.

However, our analysis had several limitations. First, the studied PAS outcomes could be more strongly associated with SARS-CoV-2 infection than with influenza infection simply because our PAS code list was assembled in part from findings of prior studies of post-COVID-19 conditions.^2,16–19^ Second, our analysis was limited to adults. Children also experience PAS associated with SARS-CoV-2^33^ and other respiratory viruses,^34,35^ suggesting similar studies should be undertaken in pediatric cohorts. Third, testing practices for SARS-CoV-2 and influenza differed during the study period. Whereas SARS-CoV-2 testing was widely undertaken for both inpatients and in ambulatory settings, influenza testing was less widely available and may have been reserved for individuals with more severe illness, or those receiving an initial negative SARS-CoV-2 test result. Our analyses adjusted for measured differences between COVID-19 cases and influenza cases; however, unmeasured sources of confounding and selection bias may remain. Fourth, as our study was restricted to individuals who received ARI diagnoses, our findings did not address PAS potentially associated with asymptomatic SARS-CoV-2 and influenza infections. Last, our analyses did not address differences in PAS risk beyond 180 days after cases’ index dates, limiting our ability to compare differences in the lengths of time COVID-19 cases and influenza cases may be at risk for PAS.

We found that PAS outcomes occurred with similar frequency among COVID-19 cases and influenza cases, although PAS among COVID-19 cases were more likely to require hospital admission than PAS among influenza cases. Whereas PAS are widely known to occur after COVID-19, our results suggest that the risk of PAS associated with influenza may be under-appreciated and worthy of further study. For COVID-19, rigorous estimates of the burden of PAS are necessary for the contemporary context of circulating variants and population immunity; such analyses should adjust for counterfactual risk absent infection through control-group comparisons.^36^ Similar estimates are also needed for influenza and other respiratory viruses, and can collectively inform the value of interventions aiming to prevent or mitigate the severity of viral respiratory infections.

## Data Availability

The raw data used in this manuscript include protected health information (e.g. dates of diagnoses and testing) that cannot be shared openly without appropriate human subjects approval and data use agreements. Kaiser Permanente Southern California (KPSC) institutional policy requires a data transfer agreement to be executed between KPSC and the individual recipient entity prior to transmittal of patient-level data outside KPSC. Requests for data can be addressed to the Central Business Office of the Department of Research and Evaluation (contact via).

## Disclaimer

The findings and conclusions in this report are those of the authors and do not necessarily represent the official position of the Centers for Disease Control and Prevention (CDC). Mention of a product or company name is for identification purposes only and does not constitute endorsement by CDC.

## Contributors statement

JAL, DEM, LRF, SS, BKA, and SYT designed the study; VH and JS generated analytic datasets; JAL and VH conducted formal analyses; IACR and LSS provided project administration; JAL, DEM, and SYT wrote the first draft of the manuscript; LRF SS, RH, and BKA provided critical intellectual feedback and revisions on subsequent drafts.

---

**Table S1:**
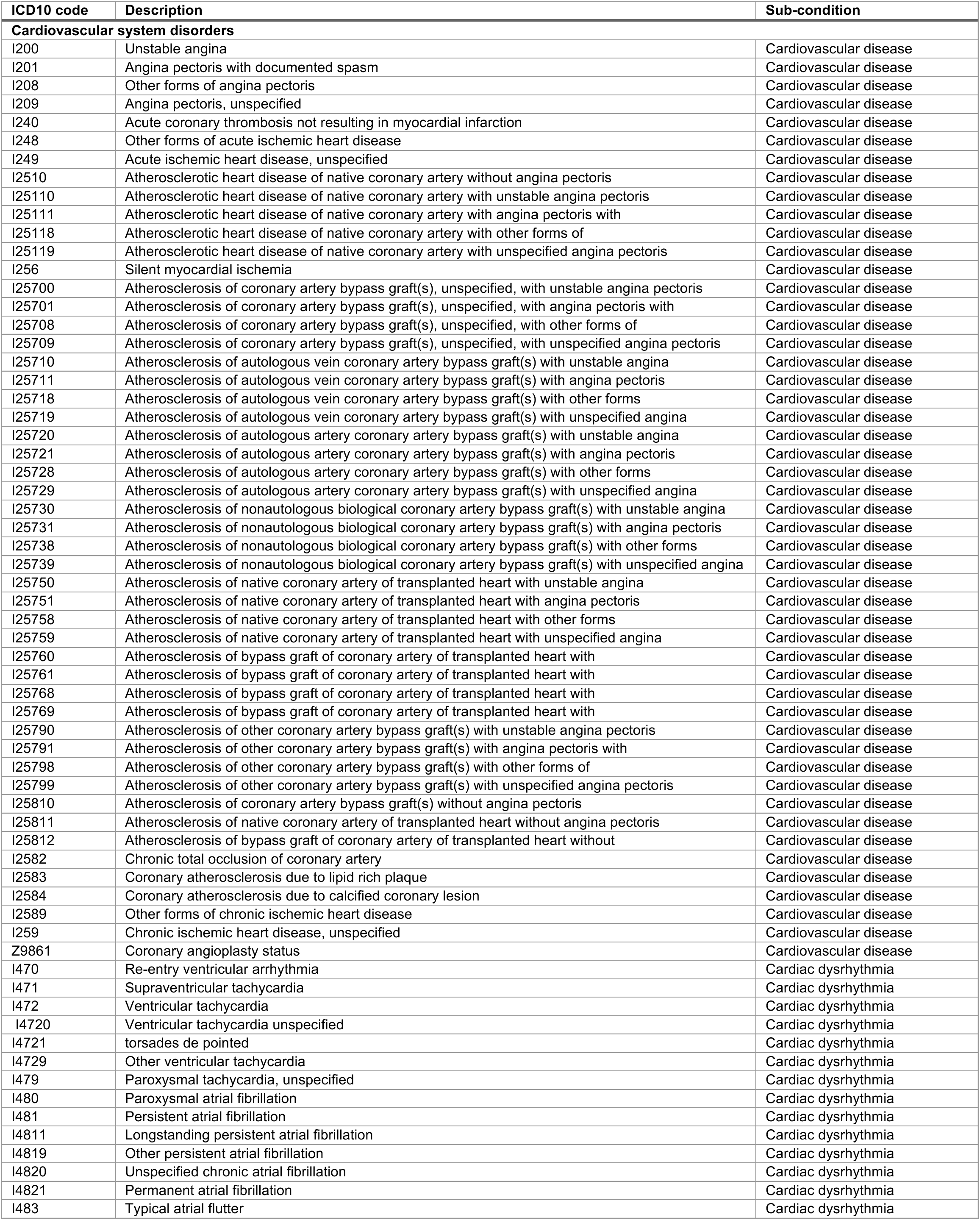
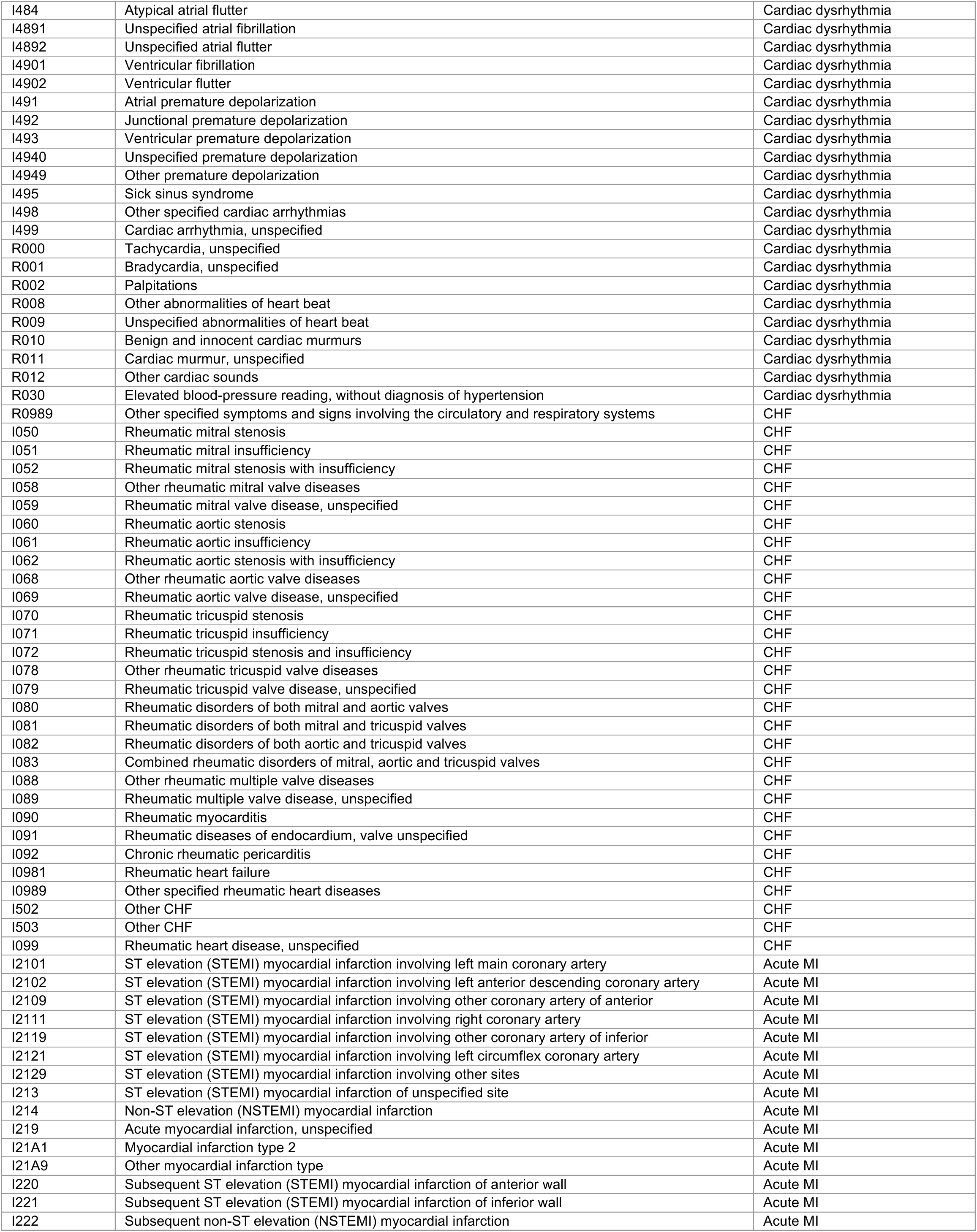
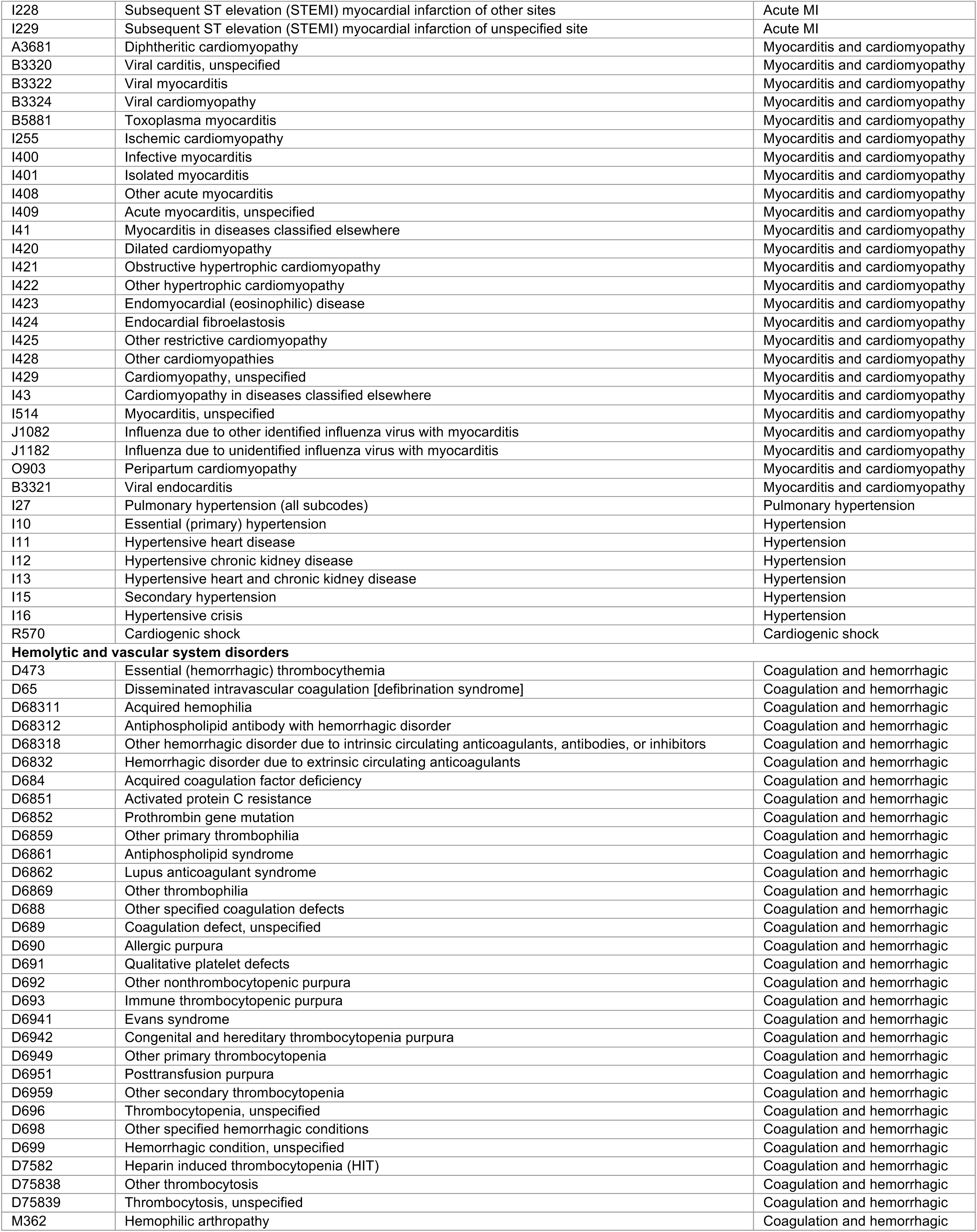
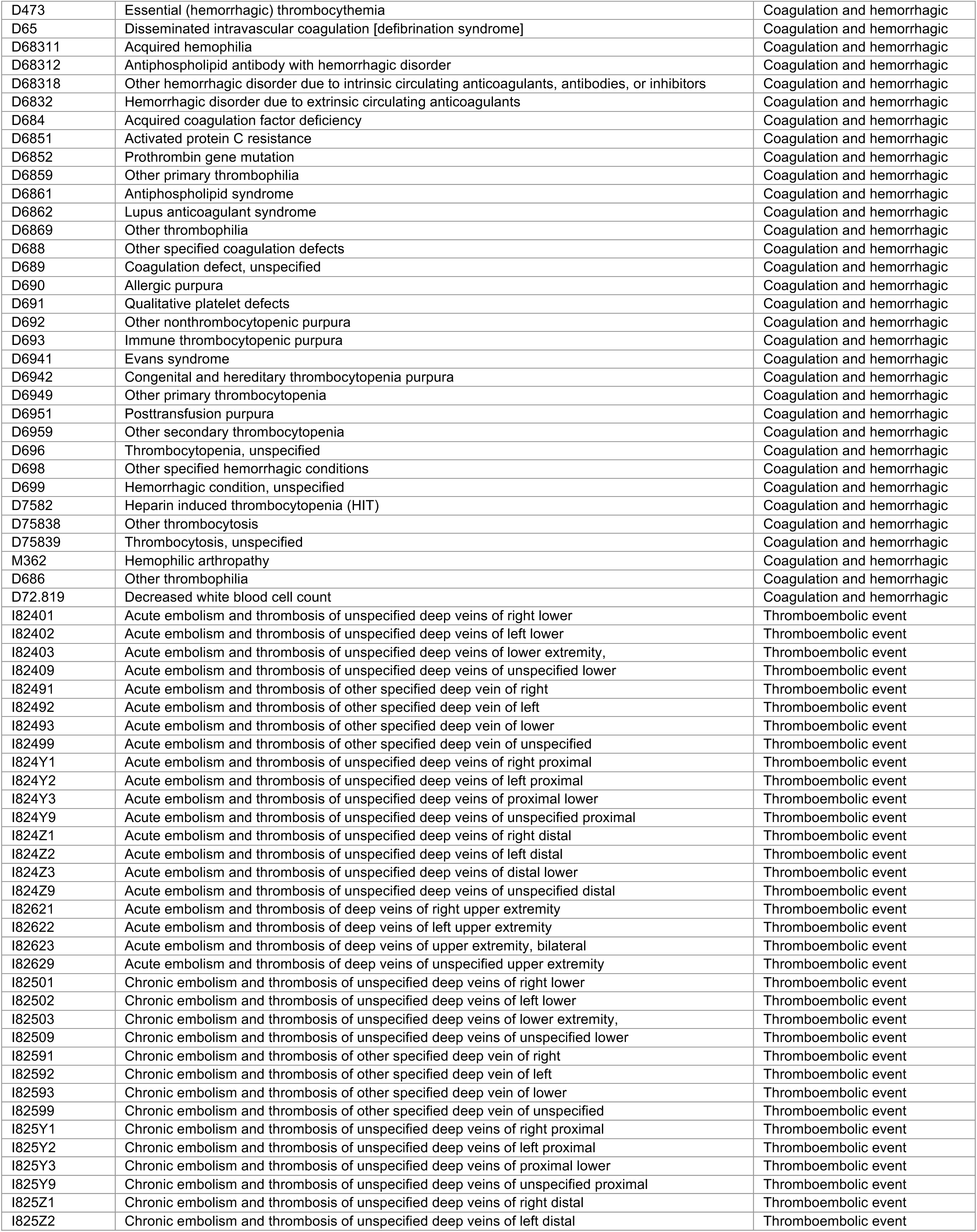
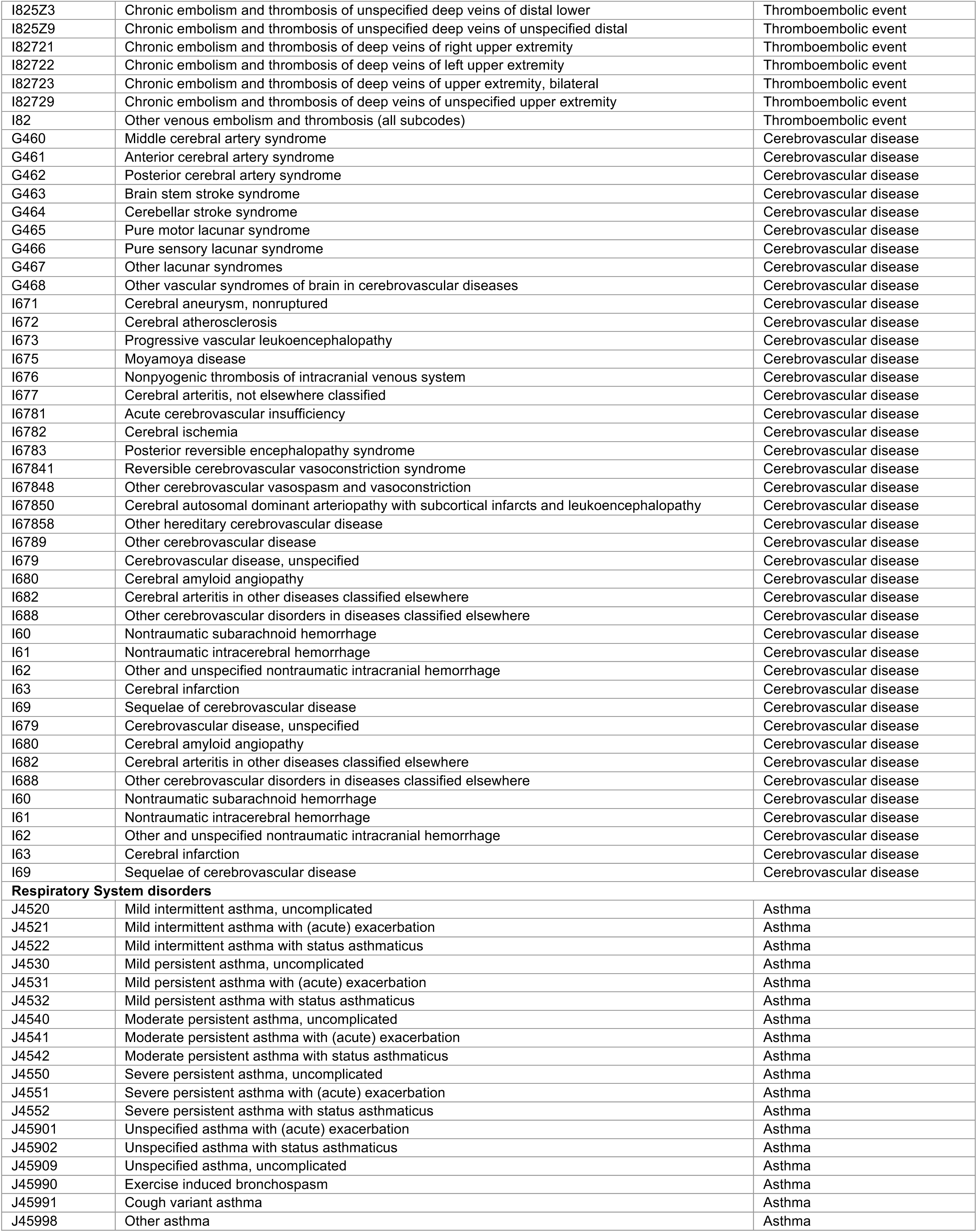
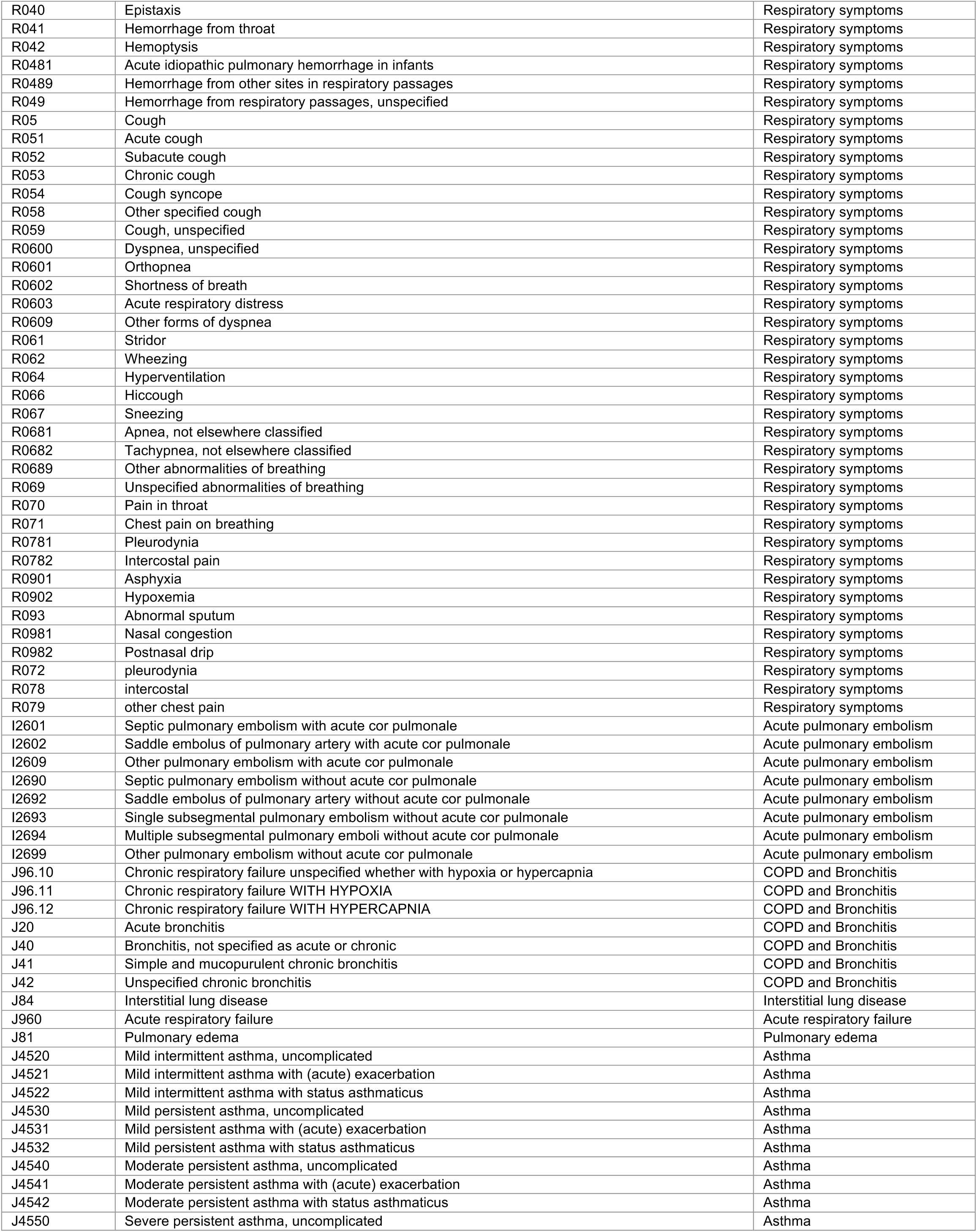
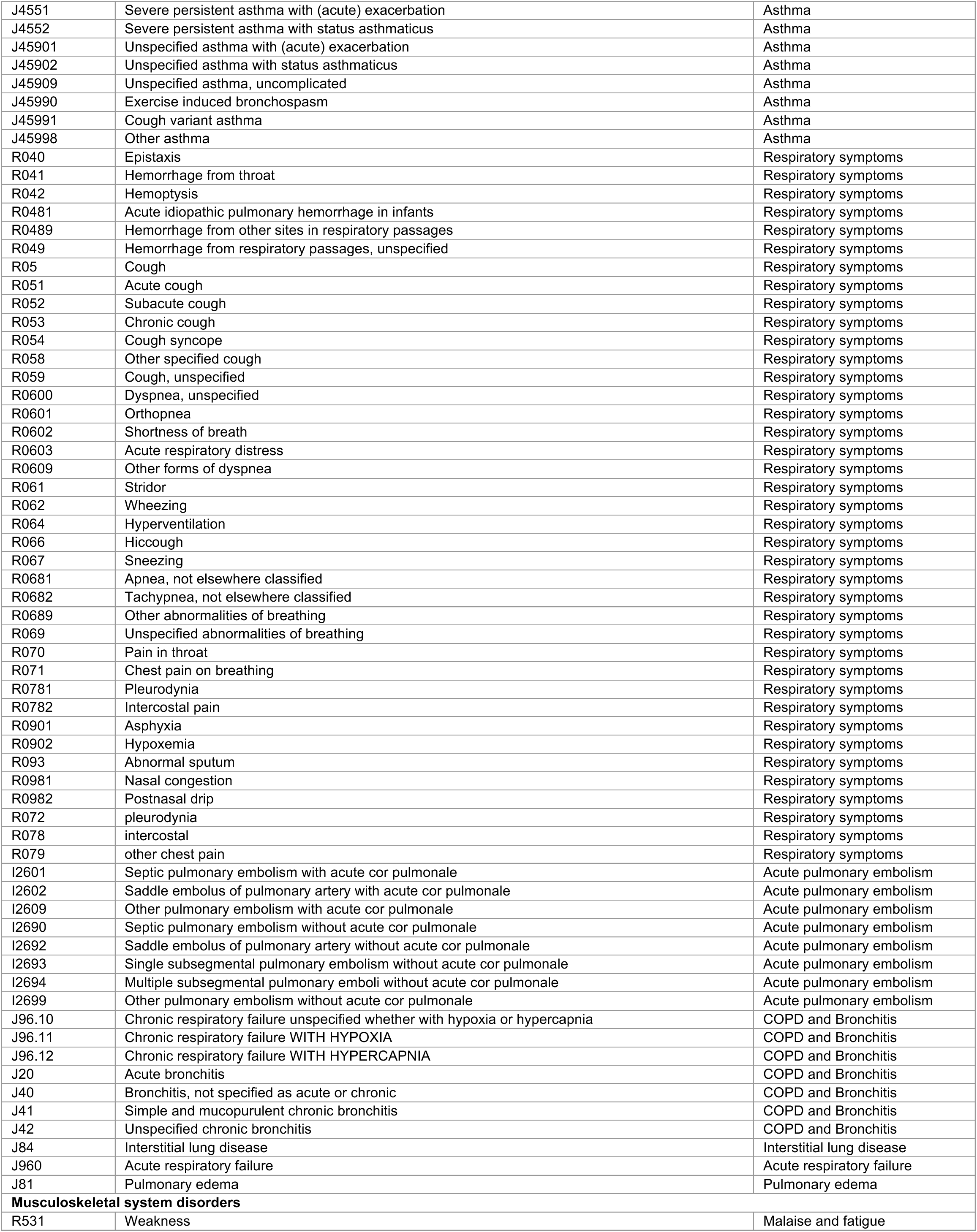
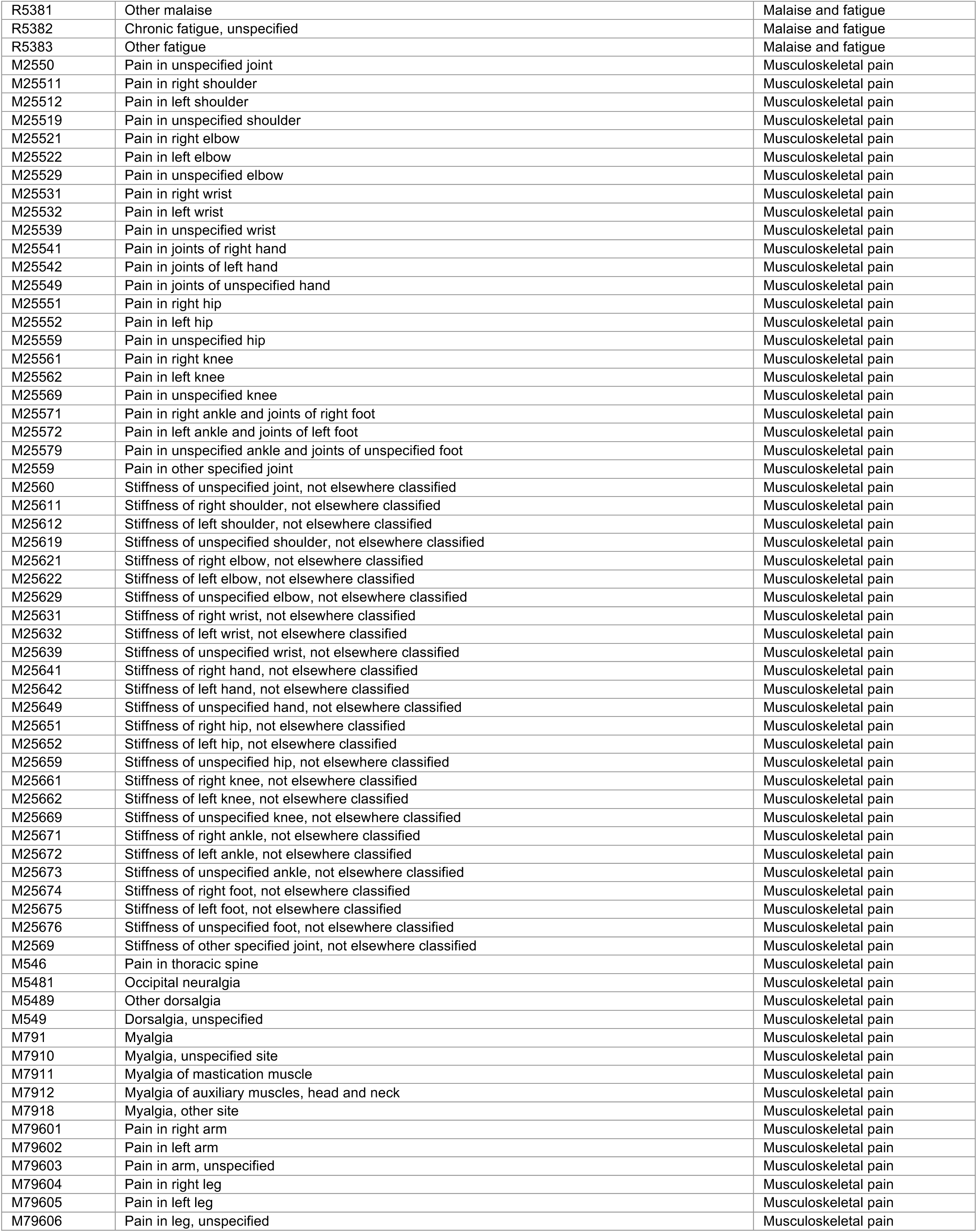
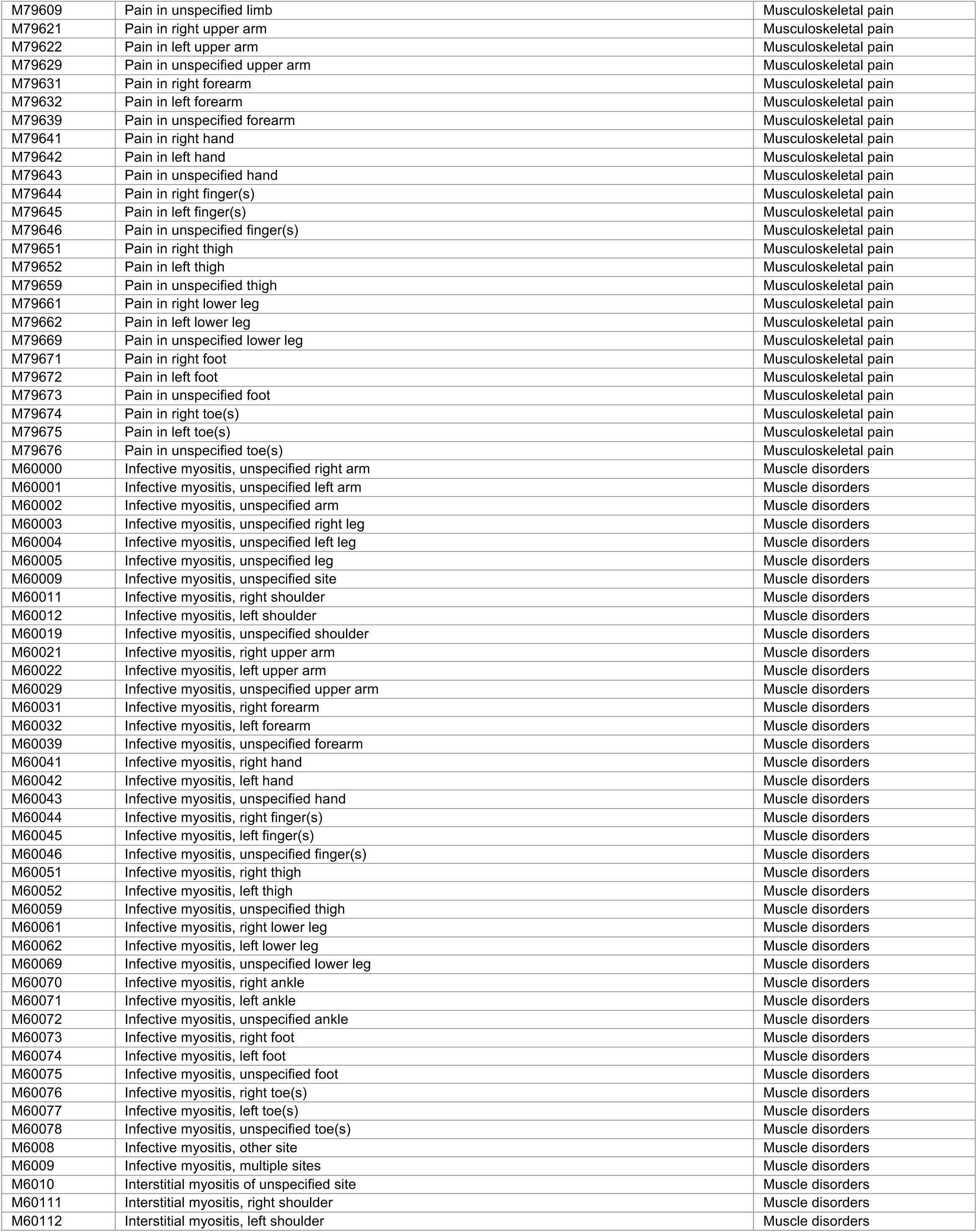
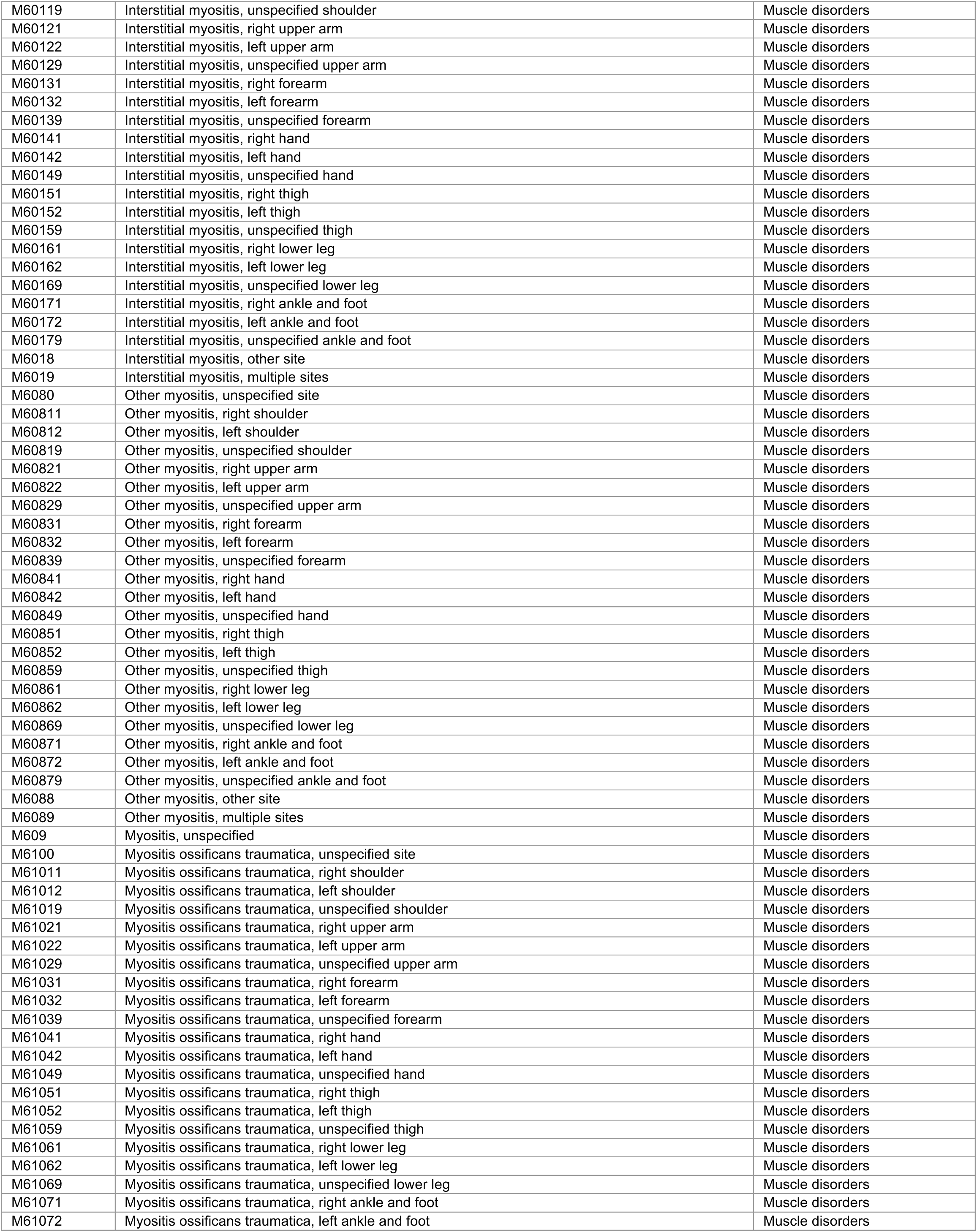
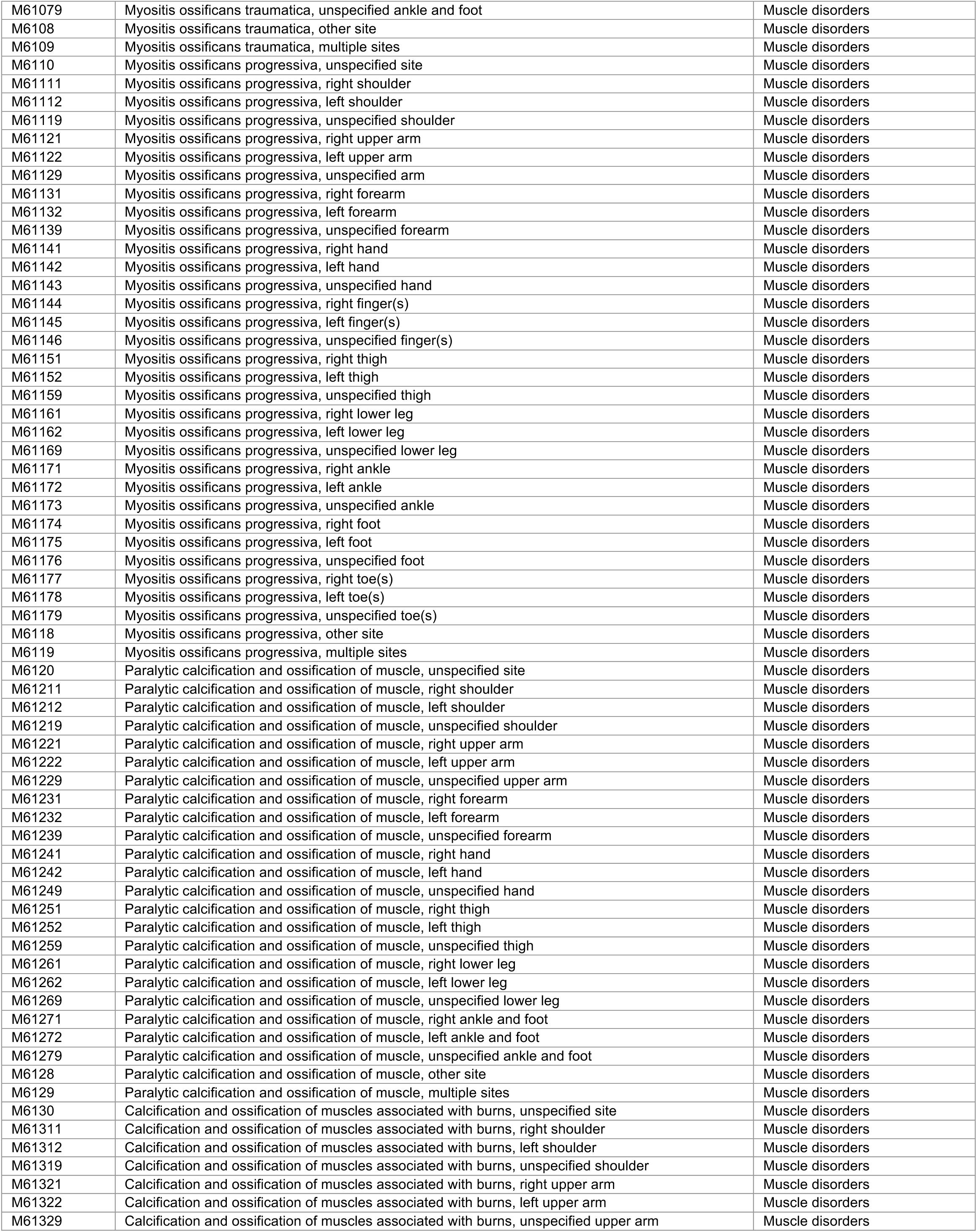
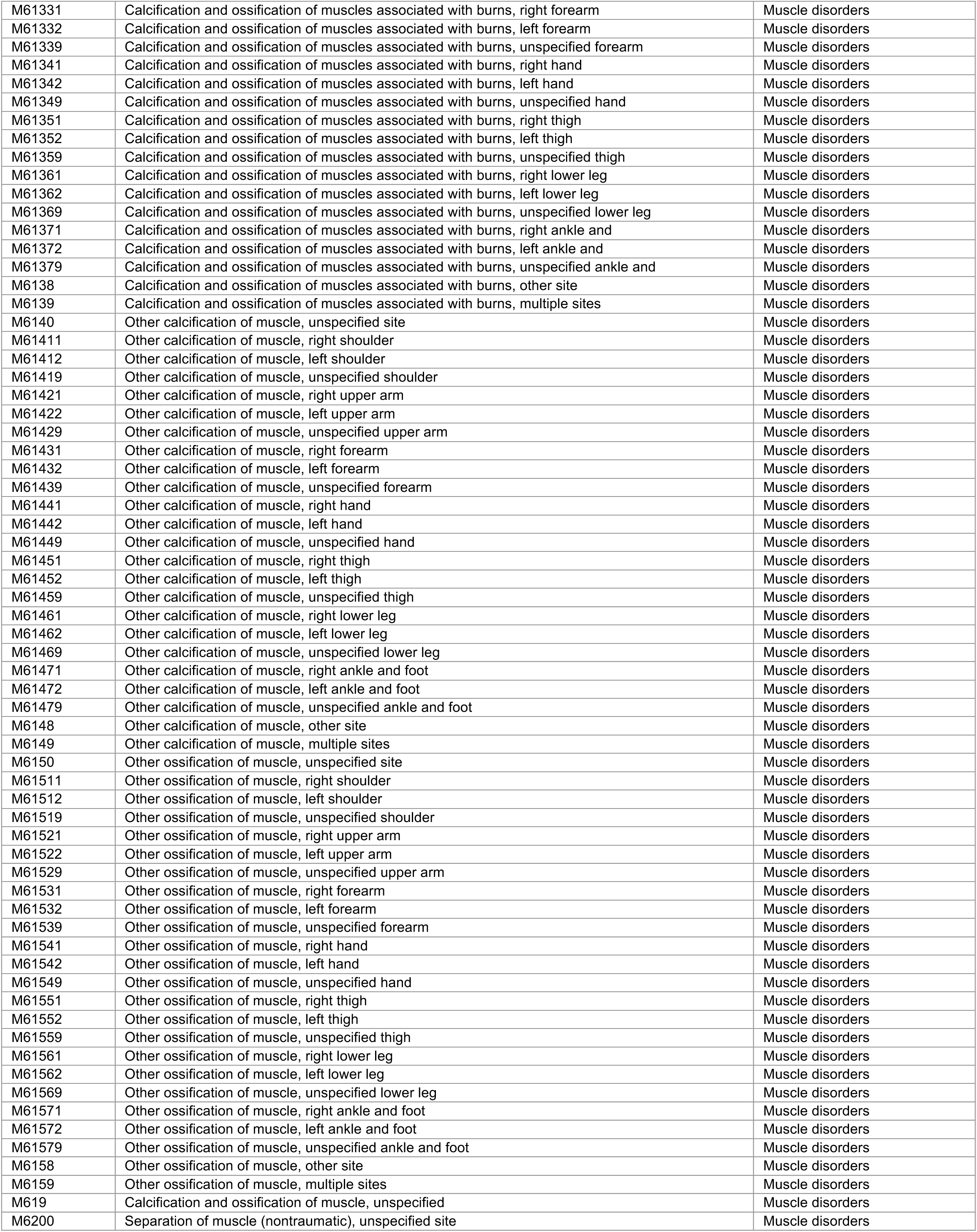
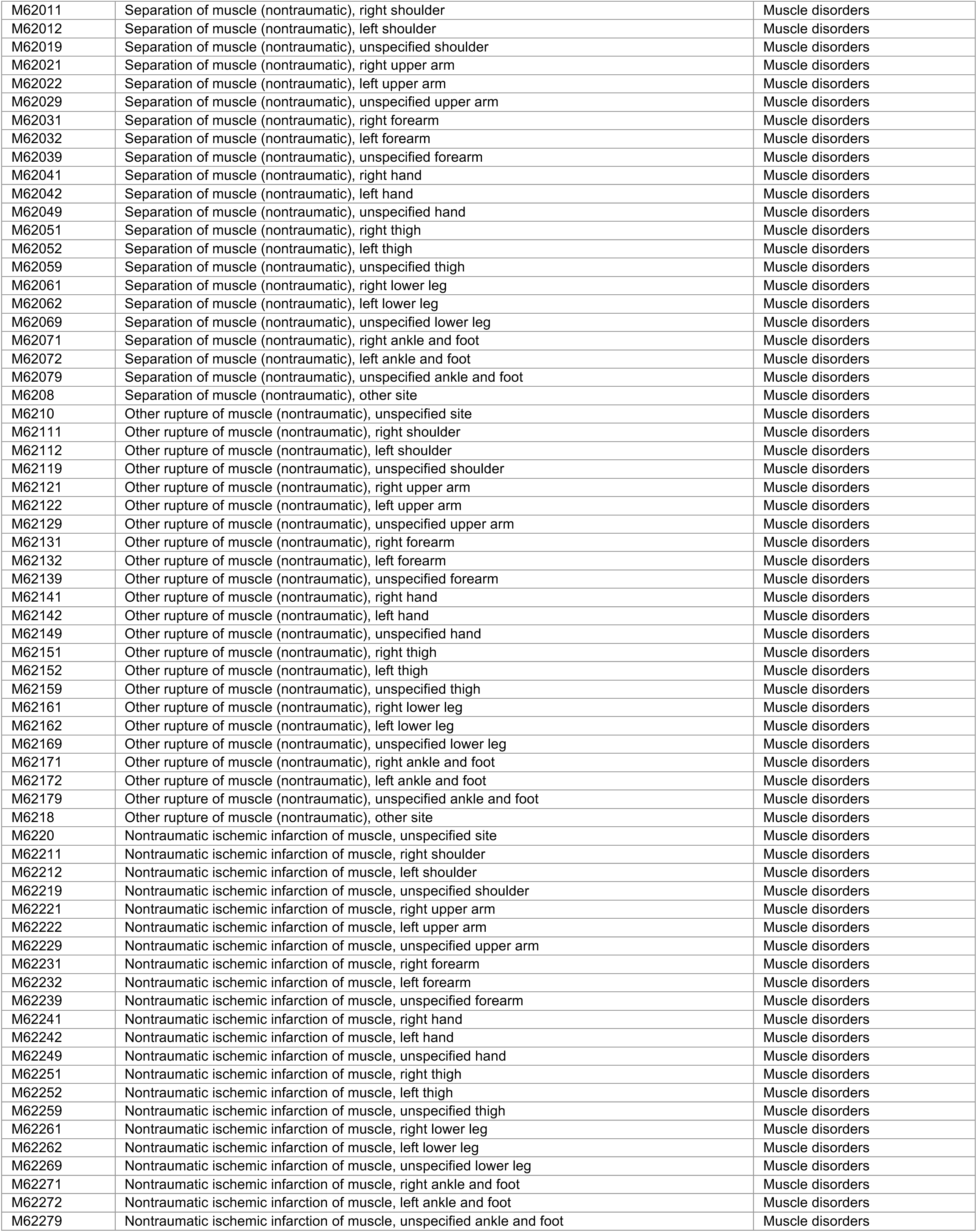
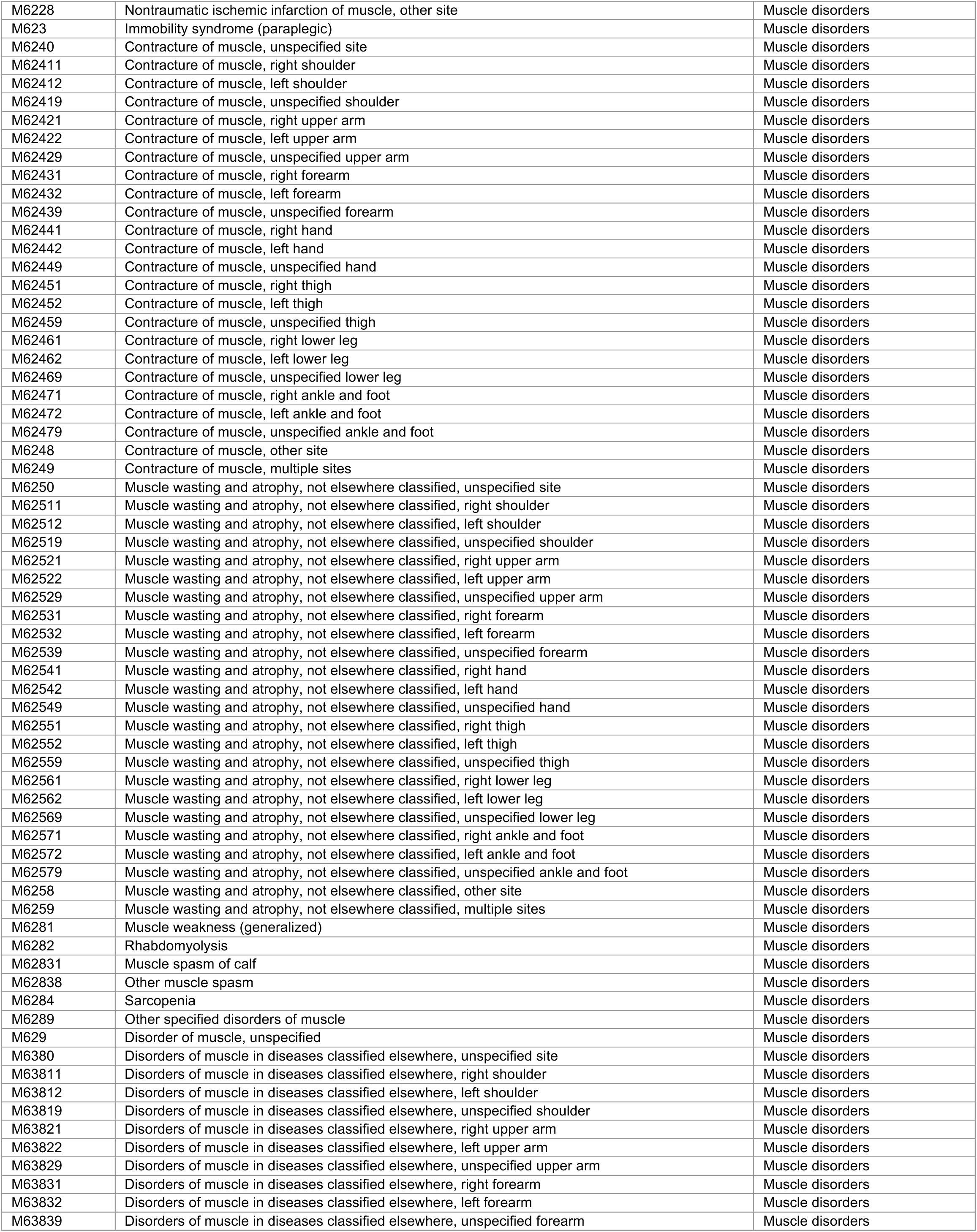
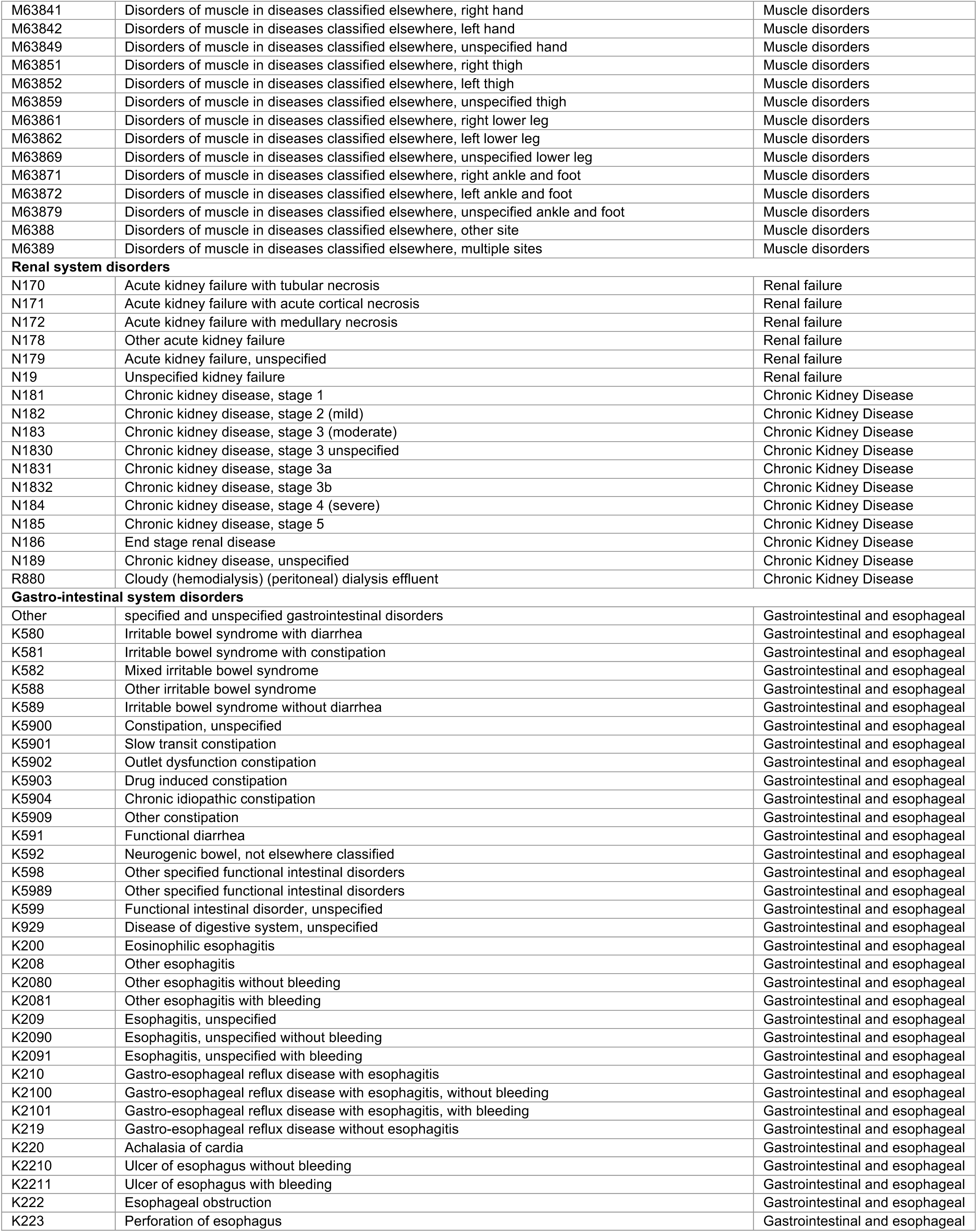
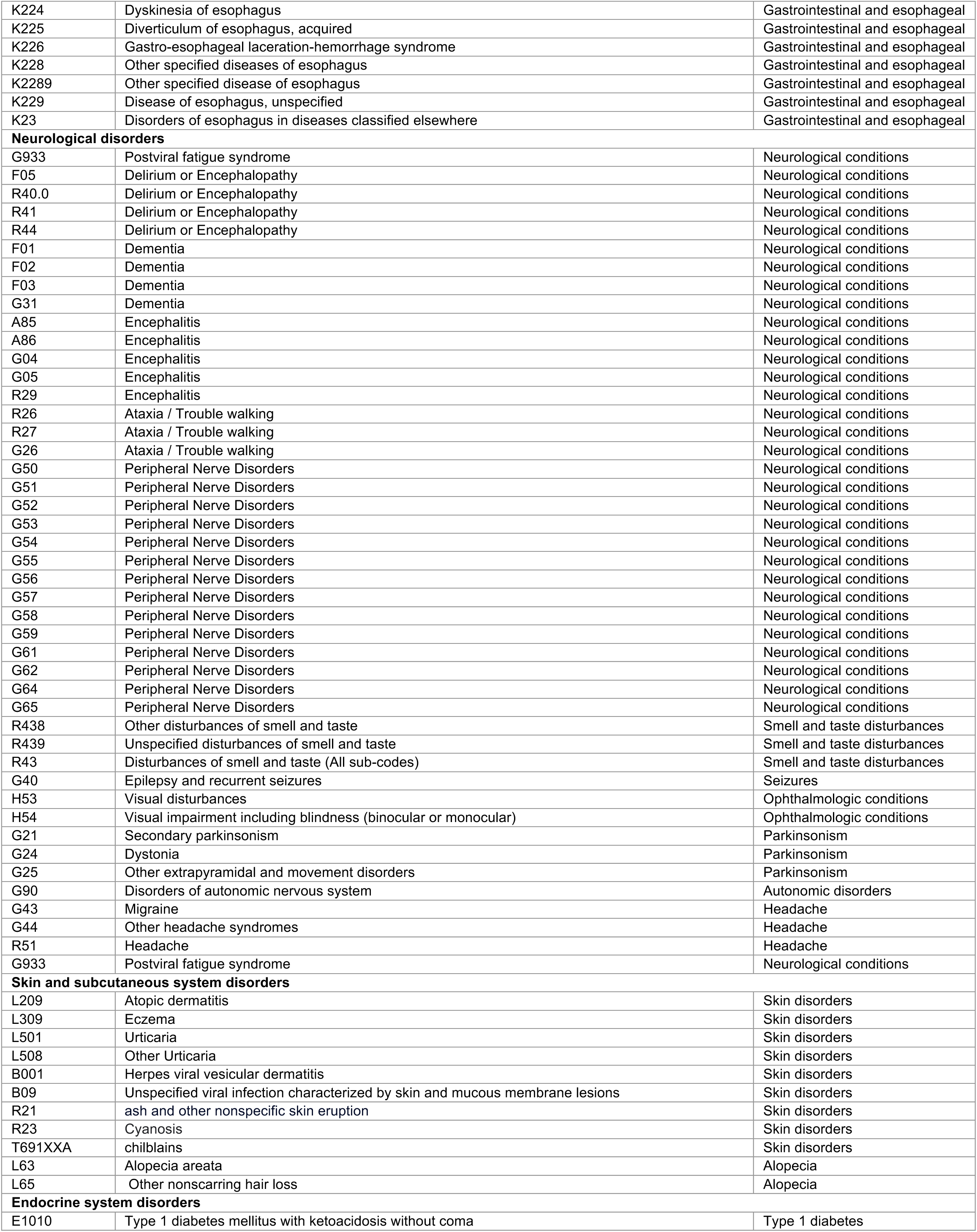
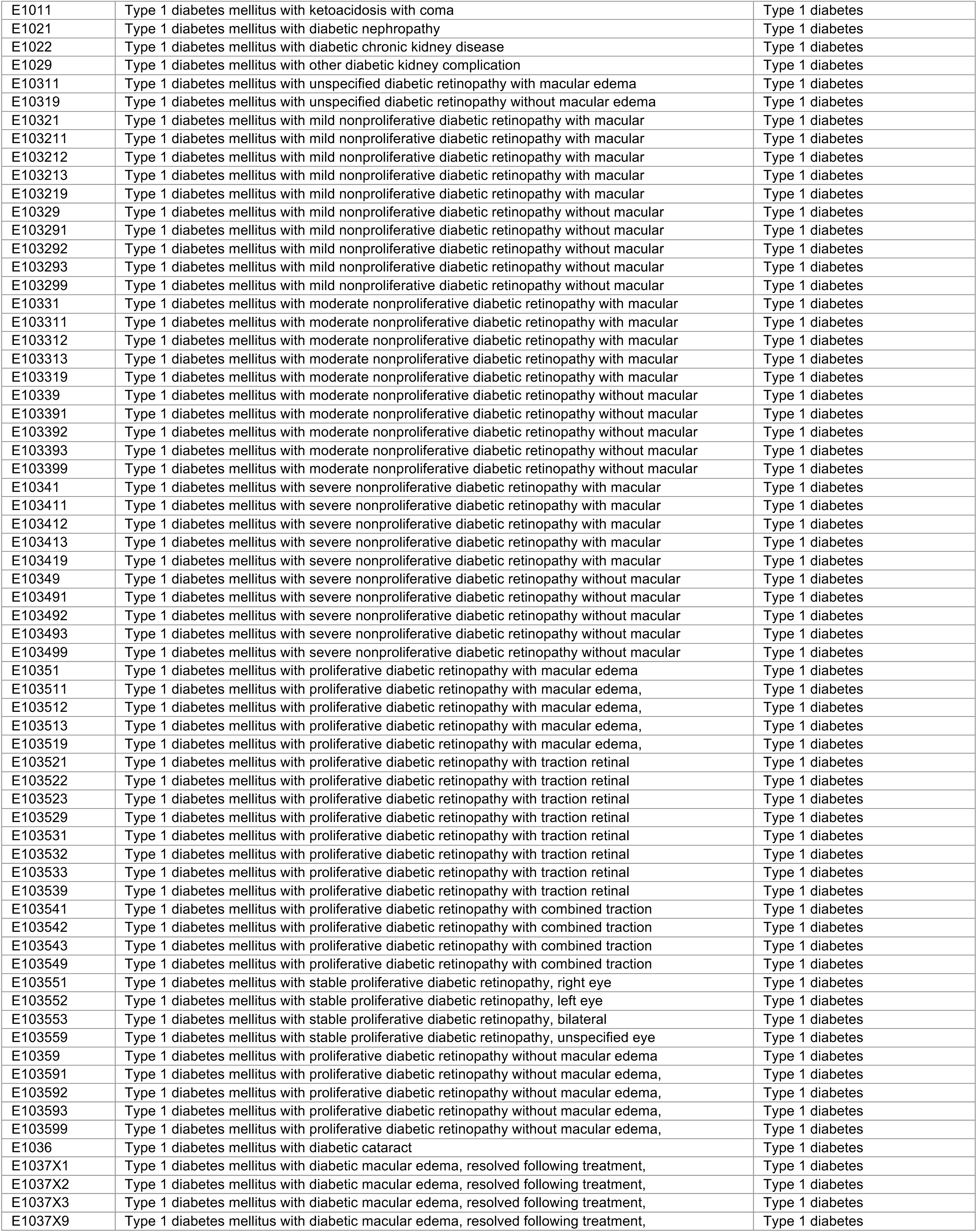
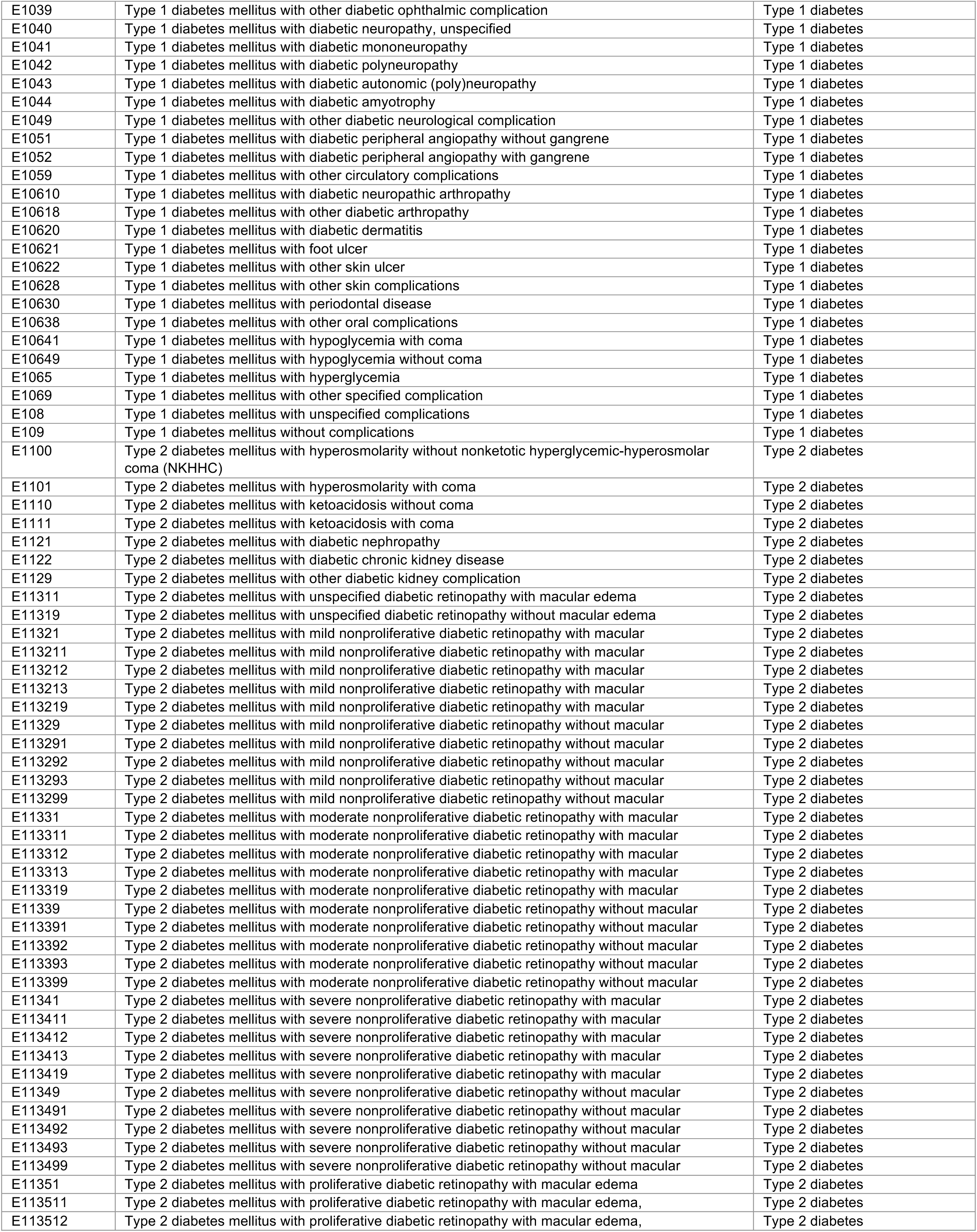
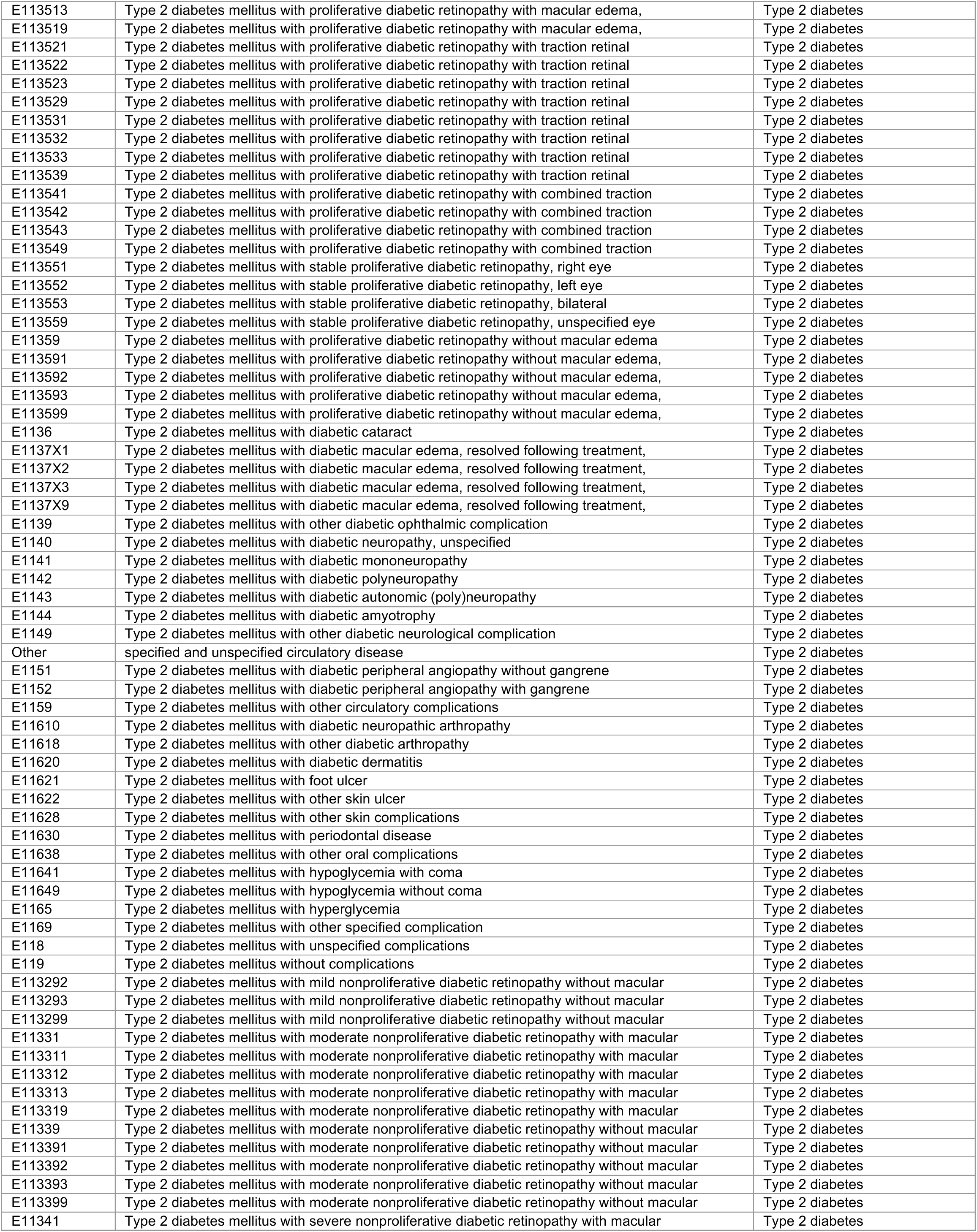
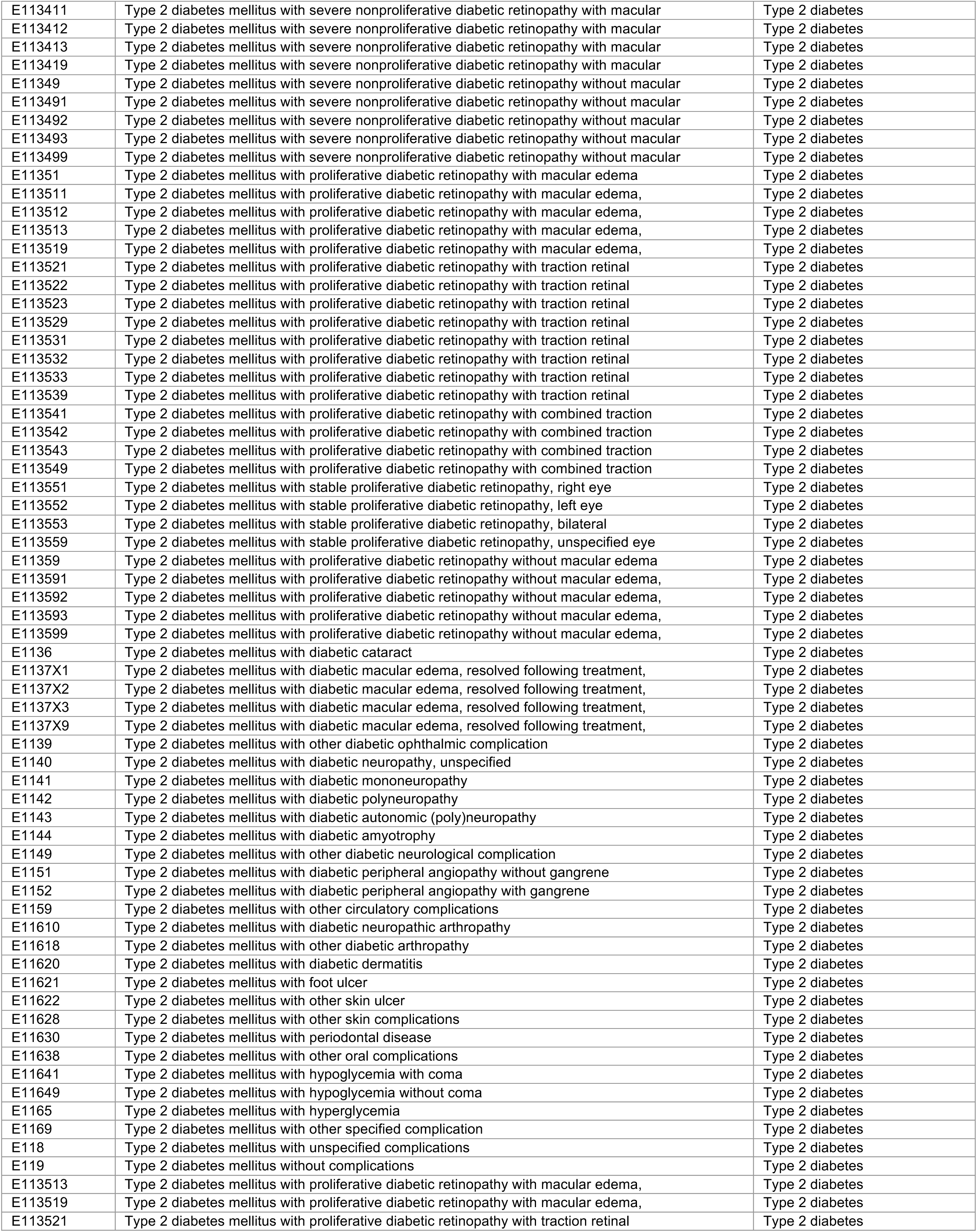
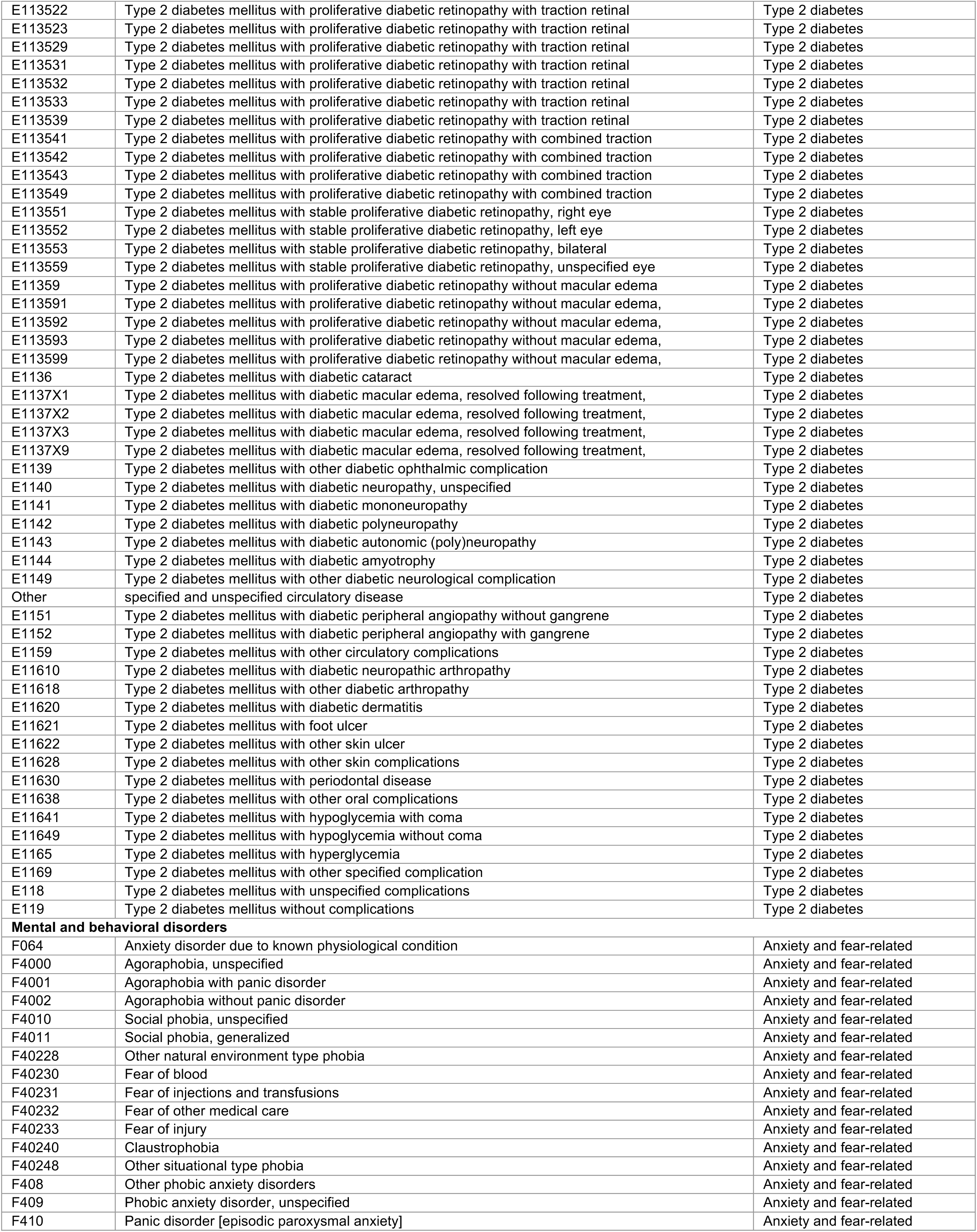
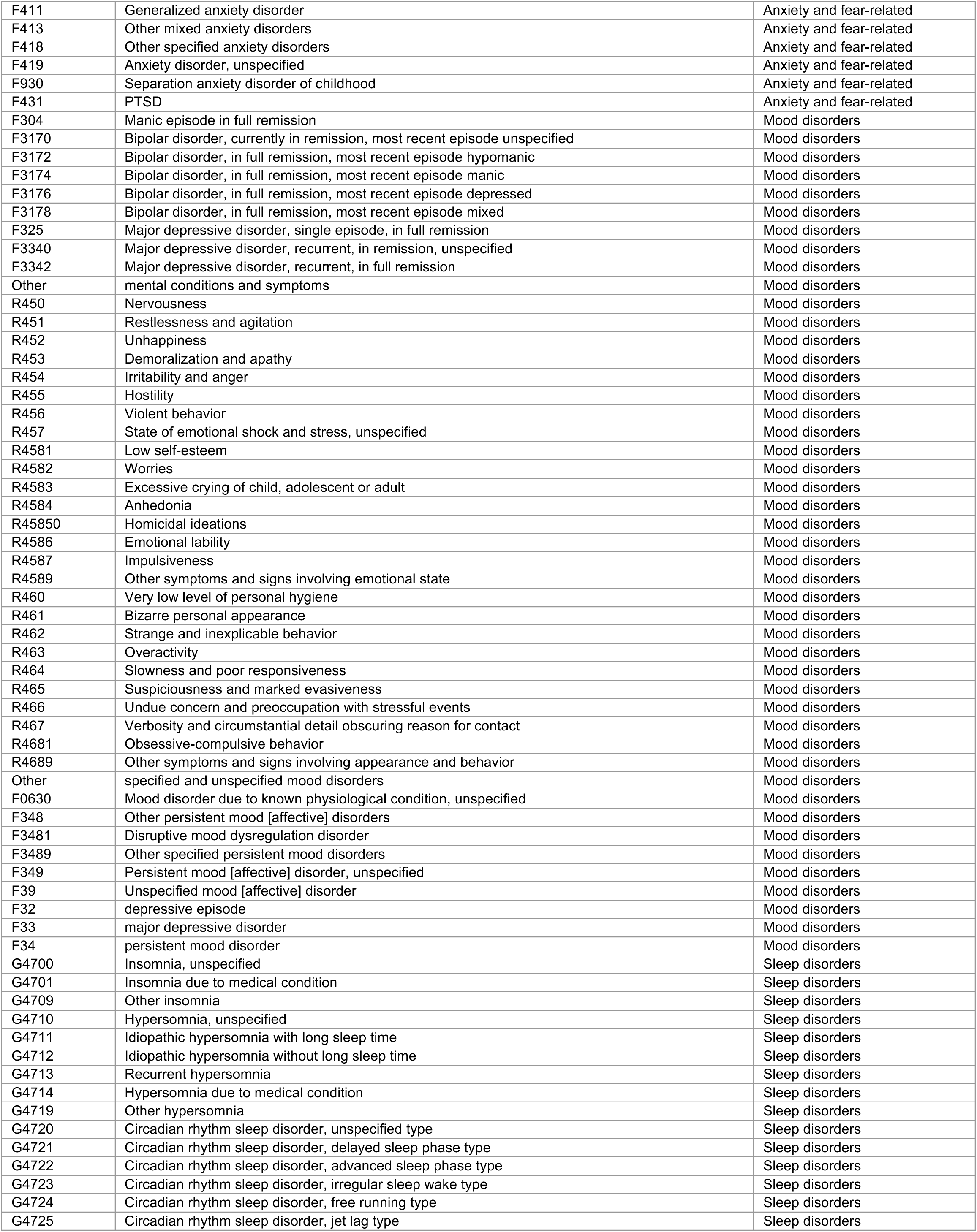
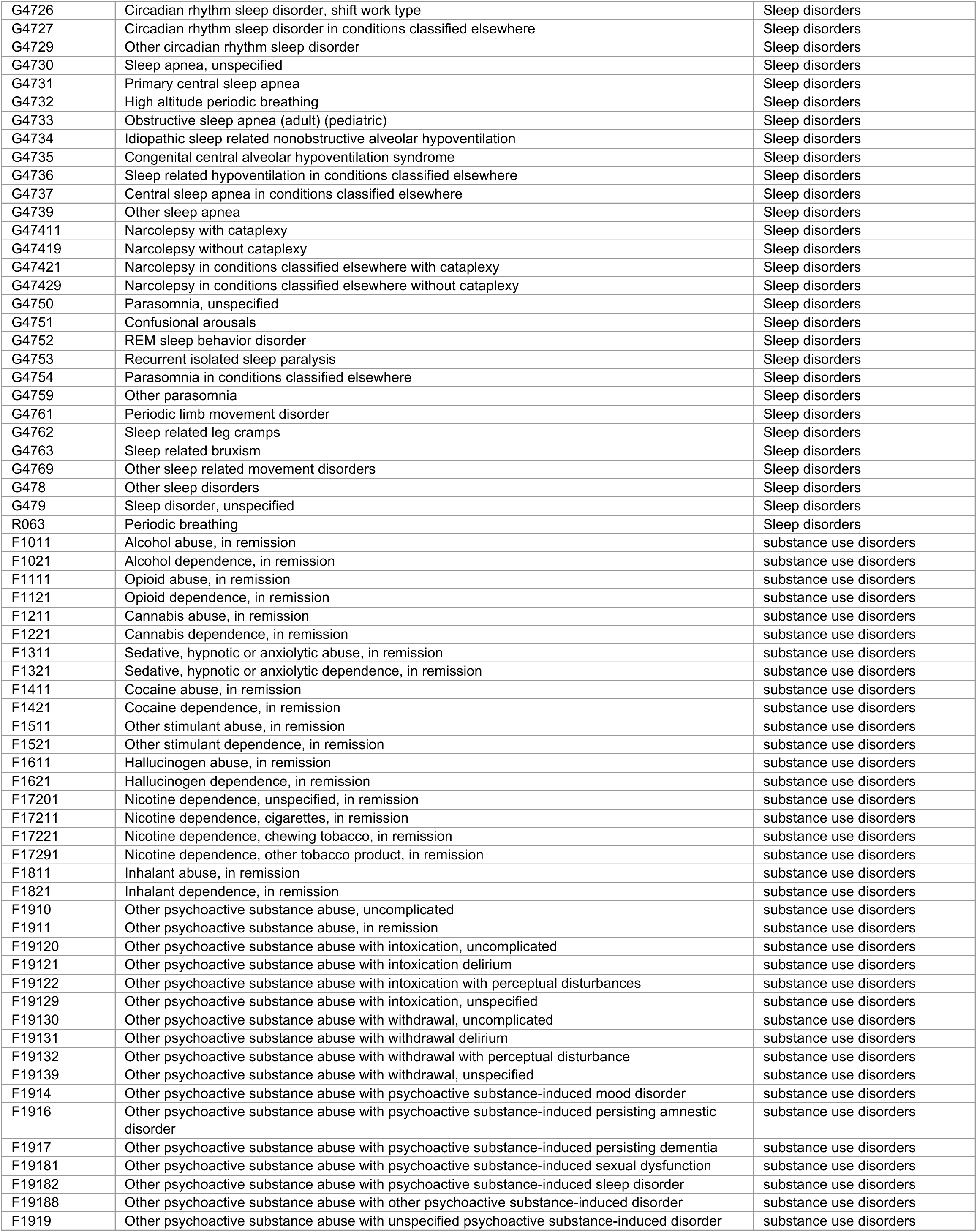
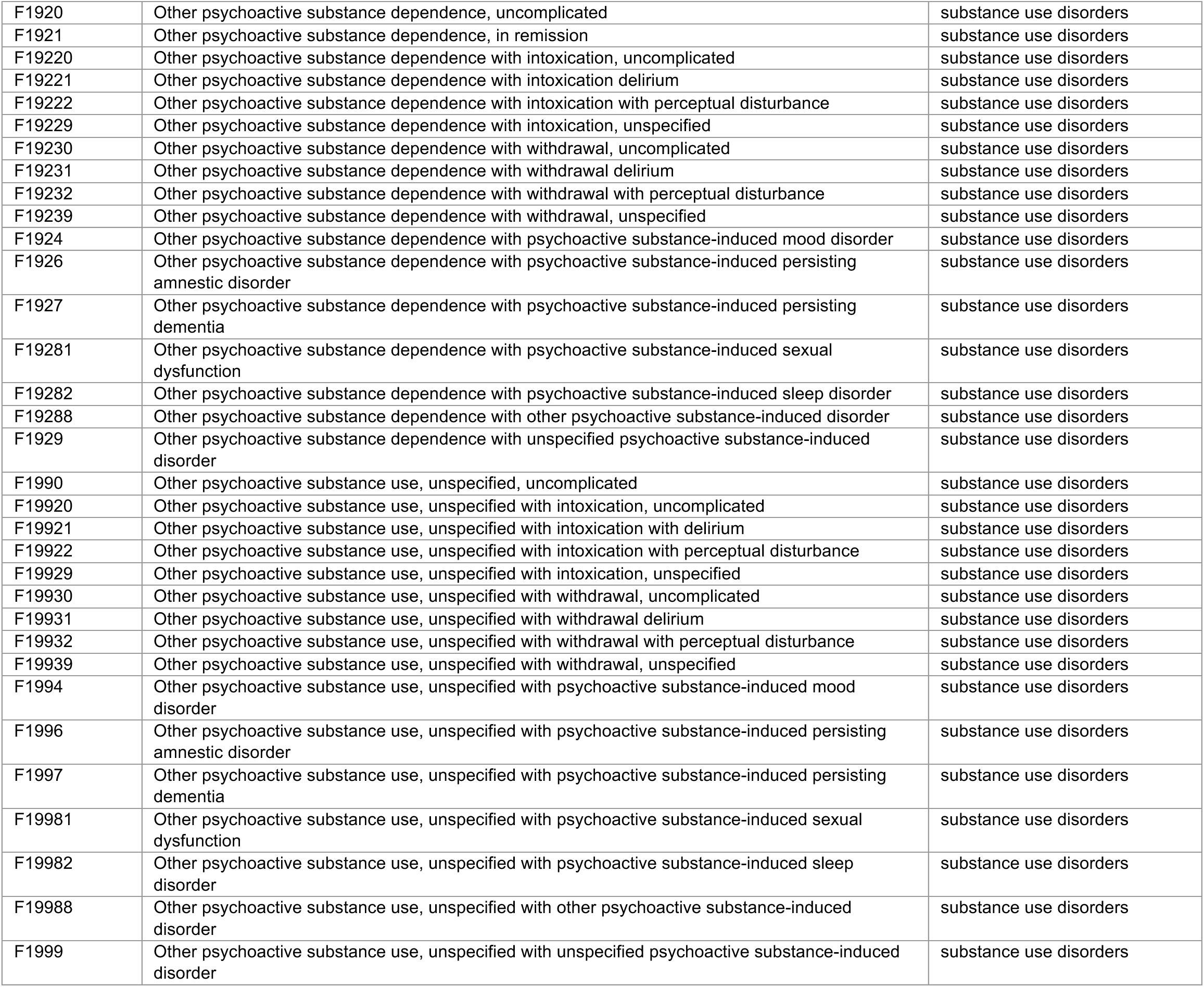
Diagnosis codes (ICD-10-CM) used to define post-acute sequelae.

**Table S2:** Cohort retention and frequency of post-acute sequelae managed in any clinical setting.

| Outcome |  | Participants, *n* (%) |  |  |  |
| --- | --- | --- | --- | --- | --- |
|  |  | Within 31-90d |  | Within 91-180d |  |
|  |  | COVID-19<br>*N*=72,745 | Influenza<br>*N*=18,395 | COVID-19<br>*N*=71,238 | Influenza<br>*N*=17,990 |
| Death/disenrollment<br>before PAS observed | Cumulatively before start of follow-up<br>period<sup>1</sup> | 1,993 (2.7% of<br>74,738) | 395 (2.1% of<br>18,790) | 3,500 (4.7% of<br>74,738) | 800 (4.3% of<br>18,790) |
|  | Deaths/disenrollments occurring during<br>follow-up period<sup>2</sup> | 973 (1.3) | 310 (1.7) | 1,108 (0.6) | 248 (1.4) |
| PAS—by category | Any | 32,878 (45.2) | 7,162 (38.9) | 37,615 (52.8) | 7,916 (44.0) |
|  | Cardiopulmonary | 14,614 (20.0) | 2,839 (15.4) | 17,134 (23.9) | 2,923 (16.1) |
|  | Hemolytic | 2,320 (3.2) | 423 (2.3) | 2,681 (3.7) | 377 (2.1) |
|  | Respiratory | 10,076 (13.8) | 2,573 (13.9) | 12,130 (16.9) | 2,738 (15.1) |
|  | Musculoskeletal | 9,063 (12.4) | 1,730 (9.3) | 11,793 (16.4) | 2,218 (12.3) |
|  | Renal | 4,466 (6.1) | 806 (4.4) | 5,074 (7.1) | 787 (4.3) |
|  | Gastrointestinal | 4,719 (6.4) | 1,030 (5.6) | 5,804 (9.1) | 1,075 (5.9) |
|  | Neurological | 6,538 (8.9) | 1,301 (7.1) | 8,453 (11.9) | 1,492 (8.2) |
|  | Skin | 1,800 (2.5) | 443 (2.4) | 2,616 (3.6) | 592 (3.3) |
|  | Endocrine | 9,706 (13.3) | 1,843 (10.0) | 11,161 (15.6) | 1,966 (10.9) |
|  | Mental health | 9,284 (12.7) | 2,185 (11.8) | 11,150 (15.5) | 2,368 (13.1) |
| PAS—by clinical setting | Any setting | 32,878 (45.2) | 7,162 (38.9) | 37,615 (52.8) | 7,916 (44.0) |
|  | Ambulatory or higher-acuity setting | 27,082 (37.1) | 5,723 (31.0) | 32,361 (45.4) | 6,526 (36.2) |
|  | Emergency department or higher-acuity<br>setting | 9,121 (12.4) | 2,067 (11.2) | 11,771 (16.5) | 2,565 (14.2) |
|  | Inpatient setting | 2,078 (2.8) | 335 (1.8) | 2,226 (3.1) | 376 (2.1) |

PAS: post-acute sequelae.
<sup>1</sup>Figures indicate the number of all otherwise-eligible COVID-19 and influenza cases (included in Table 1) who died or disenrolled before the beginning of each follow-up period. Totals represented in column headings indicate the number of individuals who remained alive and enrolled at 31 days and 91 days after index.<sup>2</sup>Figures indicate the number of COVID-19 and influenza cases who died or disenrolled during the indicated follow-up periods, calculated as a proportion of the total of all individuals who were retained in follow-up at 31 days and 91 days after index (column heading totals).

**Table S3:** Cumulative incidence of post-acute sequelae as new-onset conditions or exacerbations of existing conditions, diagnosed in any clinical setting.

| Outcome |  | Cumulative incidence per 100 persons during follow-up period, by index infection<sup>1</sup> |  |  |  |
| --- | --- | --- | --- | --- | --- |
|  |  | *Within 31-90d* |  | *Within 91-180d* |  |
|  |  | <u>COVID-19</u> | <u>Influenza</u> | <u>COVID-19</u> | <u>Influenza</u> |
| PAS—exacerbation<sup>2</sup> | Cardiopulmonary | 38.41 | 39.02 | 45.54 | 40.01 |
|  | Hemolytic | 24.46 | 29.70 | 29.84 | 23.08 |
|  | Respiratory | 21.97 | 22.77 | 26.92 | 24.32 |
|  | Musculoskeletal | 19.82 | 16.63 | 25.02 | 20.41 |
|  | Renal | 42.63 | 48.75 | 49.73 | 46.95 |
|  | Gastrointestinal | 17.81 | 18.90 | 21.83 | 18.82 |
|  | Neurological | 21.15 | 19.24 | 26.97 | 21.14 |
|  | Skin | 6.76 | 7.20 | 9.14 | 8.39 |
|  | Endocrine | 51.81 | 54.18 | 60.66 | 57.78 |
|  | Mental health | 30.66 | 32.28 | 36.16 | 33.56 |
| PAS—new onset<sup>3</sup> | Cardiopulmonary | 4.20 | 3.24 | 5.53 | 3.89 |
|  | Hemolytic | 0.94 | 0.60 | 1.09 | 0.81 |
|  | Respiratory | 7.34 | 6.96 | 9.12 | 7.90 |
|  | Musculoskeletal | 6.59 | 5.16 | 9.82 | 7.57 |
|  | Renal | 0.80 | 0.48 | 1.05 | 0.66 |
|  | Gastrointestinal | 2.92 | 2.58 | 3.87 | 3.05 |
|  | Neurological | 3.86 | 3.33 | 5.56 | 4.33 |
|  | Skin | 1.80 | 1.75 | 2.80 | 2.58 |
|  | Endocrine | 0.68 | 0.49 | 0.94 | 0.76 |
|  | Mental health | 3.50 | 3.35 | 5.09 | 4.59 |

PAS: post-acute sequelae.
<sup>1</sup>We present unadjusted cumulative incidence of each PAS outcome according to individuals' history of diagnosis or lack of diagnosis within each category.
<sup>2</sup>Analyses of exacerbation outcomes are limited to individuals with a history of any preceding diagnosis (prior to their index infection) with conditions included in the indicated PAS category.
<sup>3</sup>Analyses of new-onset PAS outcomes are limited to individuals without a history of any preceding diagnosis (prior to their index infection) with conditions included in the indicated PAS category

**Table S4:**
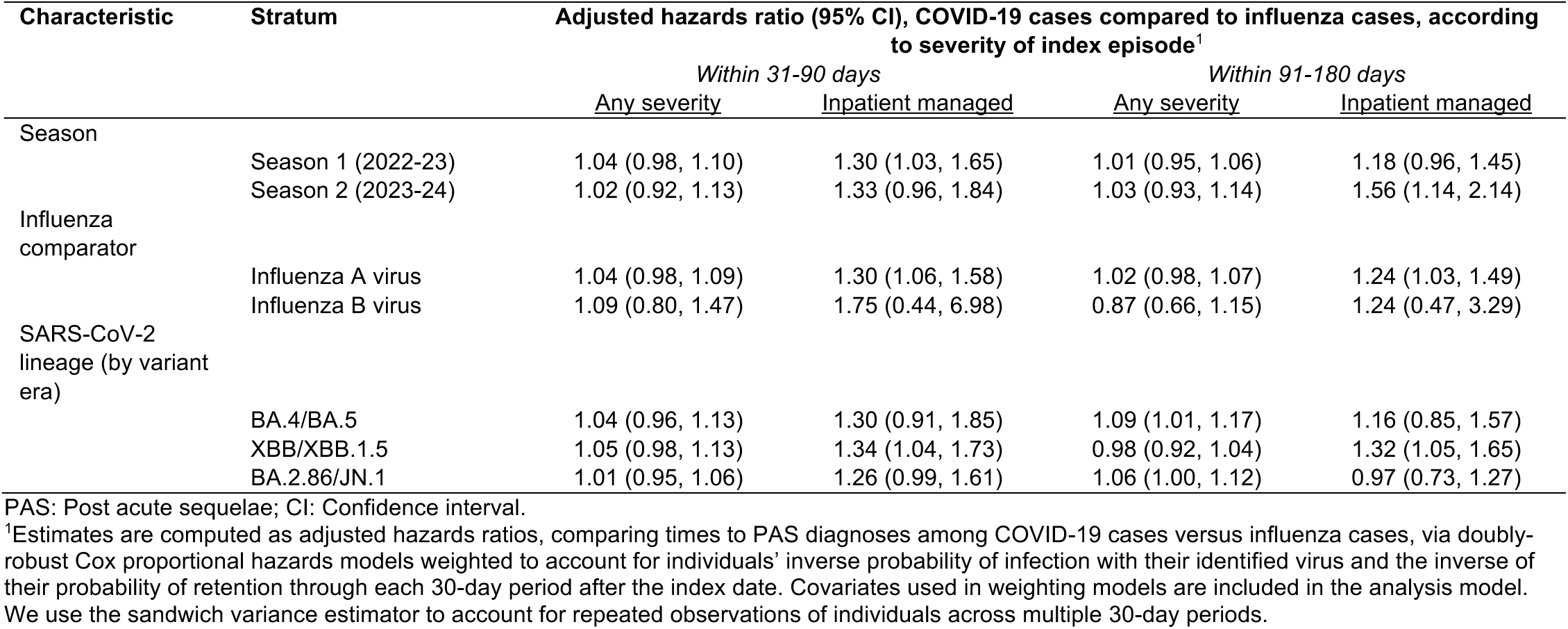
Adjusted hazard ratios of PAS outcomes, subset by infecting virus lineage or period.

